# Untargeted proteomics enables ultra-rapid variant prioritization in mitochondrial and other rare diseases

**DOI:** 10.1101/2024.08.06.24311318

**Authors:** Daniella H. Hock, Nikeisha J. Caruana, Liana N. Semcesen, Nicole J. Lake, Luke E. Formosa, Sumudu S. C. Amarasekera, Tegan Stait, Simone Tregoning, Leah E. Frajman, David R. L. Robinson, Megan Ball, Boris Reljic, Bryony Ryder, Mathew J. Wallis, Anand Vasudevan, Cara Beck, Heidi Peters, Joy Lee, Natalie B. Tan, Mary-Louise Freckmann, MitoMDT Diagnostic Network for Genomics and Omics, Vasiliki Karlaftis, Chantal Attard, Paul Monagle, Amanda Samarasinghe, Rosie Brown, Weimin Bi, Monkol Lek, Robert McFarland, Robert W. Taylor, Michael T. Ryan, Zornitza Stark, John Christodoulou, Alison G. Compton, David R. Thorburn, David A. Stroud

**Author notes:** To whom correspondence should be addressed. (Daniella H. Hock) (David R. Thorburn); (David A. Stroud). MitoMDT Diagnostic Network for Genomics and Omics members are listed at the end of the manuscript.

## Abstract

Only half of individuals with suspected rare diseases receive a definitive genetic diagnosis following genomic testing. A genetic diagnosis allows access to appropriate patient care and reduces the number of potentially unnecessary interventions and related healthcare costs. Here, we demonstrate that an untargeted quantitative mass-spectrometry approach quantifying >6,000 proteins in primary fibroblasts representing >80% of known mitochondrial disease genes can provide functional evidence for 83% of individuals in a cohort of known primary mitochondrial diseases. We profiled >90 individuals, including 28 with confirmed disease and diagnosed 6 individuals with variants in both nuclear and mitochondrial genes. Lastly, we developed an ultra-rapid proteomics pipeline using minimally invasive peripheral blood mononuclear cells to support upgrade of variant pathogenicity in as little as 54 hours in critically ill infants with suspected mitochondrial disorders. This study supports the integration of a single untargeted proteomics test into routine diagnostic practice for the diagnosis of rare genetic disorders in clinically actionable timelines, offering a paradigm shift for the functional validation of genetic variants.

## Introduction

Despite advances in genomic sequencing approaches, only 35-70% of individuals with suspected rare disease receive a molecular diagnosis following whole exome sequencing (WES) or whole genome sequencing (WGS)^1–5^. Negative cases generally remain undiagnosed due to variants being refractory to short-read sequencing approaches or when detected variants of uncertain significance (VUS, Class 3) are detected, requiring functional evidence to be upgraded to likely pathogenic (Class 4) or pathogenic (Class 5). Mitochondrial disease is an umbrella term for a group of over 300 rare monogenic disorders affecting mitochondrial energy production in the form of ATP^6^. These disorders can arise from sporadic or inherited variants in either nuclear or mitochondrial DNA (mtDNA), presenting at any stage of life with a myriad of symptoms affecting either a single organ or in a multisystemic manner^7^. Mitochondrial diseases affect approximately 1 in 5,000 live births^8^ with limited treatments available, reinforcing the importance of a genomic diagnosis for early intervention in affected individuals. A genomic diagnosis also facilitates informed reproductive options such as prenatal diagnosis (PND) and assisted reproductive technologies such as preimplantation genetic testing (PGT) and mitochondrial replacement therapy (MRT)^9^.

Functional approaches to assess VUS pathogenicity in mitochondrial disease have historically relied on targeted and low throughput tests such as respiratory chain enzymology (RCE), SDS-polyacrylamide gel electrophoresis (PAGE) or Blue Native (BN)-PAGE and immunoblotting. RCE assesses the activity of the mitochondrial respiratory chain complexes I-IV and is typically normalised to the activity of a single enzyme, citrate synthase (CS), to account for variability in mitochondrial content determined by sample quality, amount and storage^10^. Western blotting has also been used for the confirmation of VUS pathogenicity based on protein abundance or differential size, although it relies on having a strong genetic lead for disease causation from genomic data and the commercial availability of antibodies for the desired protein.

Other non-targeted functional approaches such as transcriptomics and proteomics have also been applied to the diagnosis of mitochondrial diseases. In selected cohorts, transcriptomics has been shown to increase the diagnosis of mitochondrial disorders by 10-16%^11, 12^ and other rare Mendelian disorders by 7-17%^13–15^. Although transcriptomic analysis can be a powerful tool in assessing pathogenicity of intronic and splice variants, this approach has limited power in offering functional information on missense variants, one of the most common and challenging classes of variants to assess^16, 17^. Splice variants are predicted to account for only 10% of pathogenic variants associated with autosomal recessive disorders^18^, in line with current diagnostic rates achieved by transcriptomics. Missense variants, on the other hand, account for 60% of pathogenic variants^18^ and approximately 40% of them are expected to result in reduced protein levels due to protein instability and turnover^19^. Proteomics can detect such changes and has been demonstrated to contribute to the diagnosis of Mendelian disorders by providing functional evidence for not only missense variants but also splice, deep intronic and copy number variants ^11, 20–25^. Despite this, there is a lack of data to inform the general utility of proteomics in rare disease diagnoses as an untargeted approach like genomics and transcriptomics.

Here, we present a systematic analysis demonstrating the utility of proteomics in the detection of rare mitochondrial disorders and provide the first validated pipeline for ultra-rapid functional testing using peripheral blood mononuclear cells (PBMCs). Reference proteomics data and associated bioinformatics tools for performing the proteome analysis approaches developed for these investigations can be applied to user-uploaded data through our interactive web tool (https://rdmassspec.shinyapps.io/RDMSExplorer/).

## Results

To validate our untargeted proteomic approach in the detection of mitochondrial disorders, we first analysed primary fibroblast cell lines from a cohort of 24 patients with confirmed genetic diagnoses based on known pathogenic or likely pathogenic nuclear or mtDNA-encoded variants. This validation cohort (VC) included cell lines with defects in subunits or assembly factors for each of the five individual OXPHOS complexes as well as defects of mtDNA translation that impact multiple complexes, causing a combined OXPHOS biochemical defect (**Fig 1A, Supplementary Table 1**). Principal component analysis showed that proteomes segregate according to their primary defect (**Fig. 1B**) and hierarchical clustering of OXPHOS subunits and assembly factors identified clear profiles (**Supplementary Fig. 1A, B**). We have previously shown similar profiles to be present in gene-edited HEK293T knockout of Complex I subunits^26^. Comparison of mitochondrial proteomes from our validation cohort of fibroblast cell lines and corresponding gene-edited HEK293T knockout cell lines (knockout cohort, KC) showed good correlation of dysregulated proteins (**Supplementary Fig. 2A, B**).

**Figure 1.**
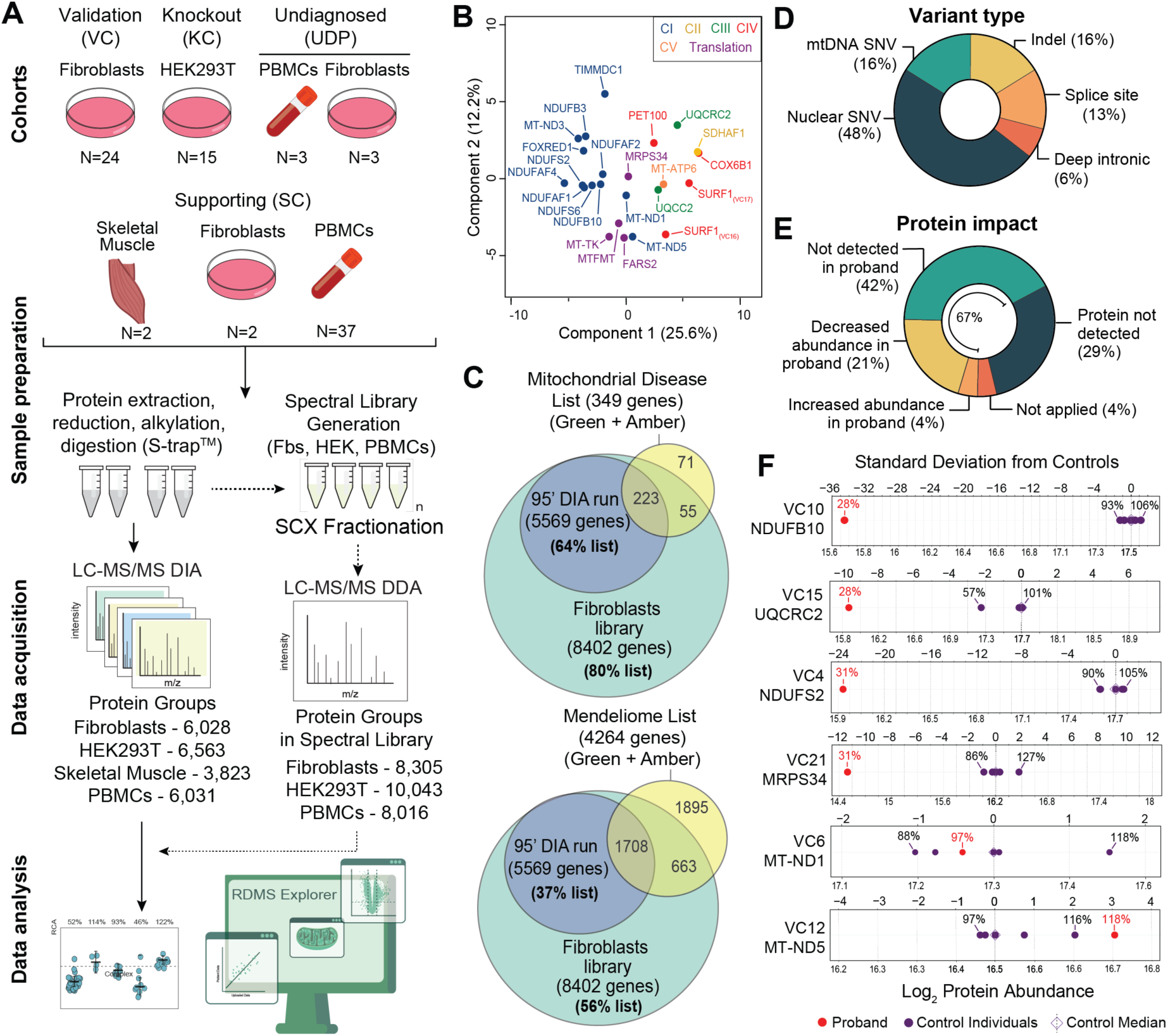
Study design and validation cohort analysis. **A –** Quantitative proteomics experimental design overview. A fibroblast validation cohort (VC), HEK293T knockout cohort (KC), undiagnosed patient cohort (UDP) and supporting cohort (SC) prepared using S-Trap^TM^ columns. Digested peptides were subjected to Liquid Chromatography tandem Mass Spectrometry (LC-MS/MS) Data Independent Acquisition (DIA). Pooled control peptides were fractionated using strong cation-exchange (SCX) chromatography and data acquired as Data Dependent Acquisition (DDA) to generate spectral libraries for fibroblasts (Fbs), HEK293T (HEK) and PBMCs. Raw data were searched using Spectronaut^®^ software and data analyses were performed with a combination of Perseus software, Python and R. **B –** Principal Component Analysis (PCA) of the fibroblast validation cohort (N=24) relative to controls based on the differential abundance of whole cell proteins calculated from t-test. **C –** Venn diagram showing the coverage of Mendeliome (v. 0.12869, top panel) and Mitochondrial disease (v. 0.787, bottom panel) lists including green (diagnostic-grade) and amber (borderline diagnostic-grade) genes retrieved from PanelApp Australia^27^. **D –** Summary of the genetic variant types analysed in the validation cohort (VC). **E –** Summary of the findings in the VC based on protein identification and abundance. In 42% (10/24) of the cases the protein expected to be affected by the genetic variant is not detected in the patient while it is detected in the controls. In 29% (7/24) of the cases, the protein is not detected via quantitative proteomics, in 21% (5/24) of the cases, the protein is decreased in abundance in the patient compared to the 5 controls, and in 4% (1/24) of the cases, the protein is increased compared to the controls analysed. SNV = single nucleotide variant. **F –** Protein standard deviation from control median of the respective affected gene in controls (purple) and probands (red) in the validation cohort (VC). Standard deviation was calculated from the median control variance.

The maximum theoretical coverage of known mitochondrial disease gene products from the fractionated library was 80%, with a routine coverage of 64% in a standard 95-minute library-supported DIA run from fibroblast samples (**Fig. 1C**). Our approach could also detect proteins corresponding to 37% of the Mendeliome (i.e., encoded by known disease genes) in a standard 95-minute run, supporting the potential utility of proteomics as a disease agnostic test in the resolution of many rare monogenic disorders. In terms of variant type in the validation cohort, the most common type was nuclear single nucleotide variants (48%) followed by mtDNA single nucleotide variants (16%), indel (16%), splice site (13%) and deep intronic (6%) (**Fig. 1D**). We quantified the abundance of the protein of interest in 67% of our investigations with the most common outcome being the protein abundance readily quantified in controls and absent in proband (42%) followed by a decrease in the patient relative to controls (21%) (**Fig. 1E**). In 29% of cases the protein of interest was not detected in either proband or control fibroblasts. For the cases where the protein was identified in both proband and control fibroblasts, analysis of the standard deviation show that ND1 abundance in VC6 (*MT-ND1*) was 97% relative to control median and within the control distribution, while ND5 abundance in VC12 (*MT-ND5*) was 118% relative to control median and approximately 3 standard deviations above the control median (**Fig. 1F**). Other identified proteins were outside the control range and >9 standard deviations below control median (**Fig. 1F**).

Mitochondrial content has previously been calculated in proteomics data using the mean abundance of experimentally validated mitochondrial proteins in the MitoCarta 3.0 database^28, 29^. We noticed that the mean abundance of mitochondrial proteins as a group varied between cell lines in our validation cohort (**Supplementary Fig. 3A**) but not control individuals (**Supplementary Fig. 3B**). This phenomenon is typically taken into consideration in enzymology analyses by expressing enzyme rates as citrate synthase (CS) ratios to account for mitochondrial proliferation or cell/tissue variability^10, 30^. CS enzyme activity in our validation cohort varied greatly (**Supplementary Fig. 3A, upper panel**), consistent with altered mitochondrial content. Surprisingly, while CS activity did have a weak positive correlation with the relative abundance of CS protein as detected by proteomics (**Supplementary Fig. 3C, left panel**), it did not correlate with mitochondrial content calculated from the abundance of mitochondrial proteins (**Supplementary Fig. 3C, middle panel**). In line with this, CS protein abundance also did not correlate with mitochondrial content calculated from proteomics data (**Supplementary Fig. 3C, right panel**). Since the nature of proteomics-based RCA is relative protein abundance, we concluded that mean mitochondrial protein abundance is the appropriate metric to normalise proteomics data. We built a differential mitochondrial abundance correction step into our RCA calculations (see **Materials and Methods** for detailed information). This correction is of benefit for cell lines that may have different mitochondrial content to controls and corrects for mitochondrial content before calculating the relative abundance of each complex.

We next sought to systematically benchmark our RCA analysis against clinical RCE. RCE was performed on 23 of the 24 fibroblast lines in the validation cohort, and was also performed on skeletal muscle (SKM) or lymphoblastoid cell lines (LCLs) for 17 of the 24 patients. Results are presented relative to CS (**Supplementary Table 2**) and the Bernier criteria^31^ were used to classify results. Fibroblast RCE detected a definite defect (<30%, major criterion) in the expected complex(es) for 19 of 24 fibroblast lines (79%) while three [VC4 *(NDUFS2),* VC7 *(NDUFAF1)* and VC10 *(NDUFB10*)] had a probable defect (minor criterion) and RCE did not detect a defect in VC19 (*COX6B1*) fibroblasts (**Fig. 2A left panel, Supplementary Table 2**). When skeletal muscle or LCLs were also available, RCE revealed a major defect in 9 of 17 of the analyses (53%), and a minor defect in four (23%) while three were not deficient (18%) and one was not available (6%)(**Fig. 2A, right panel)**.

**Figure 2.**
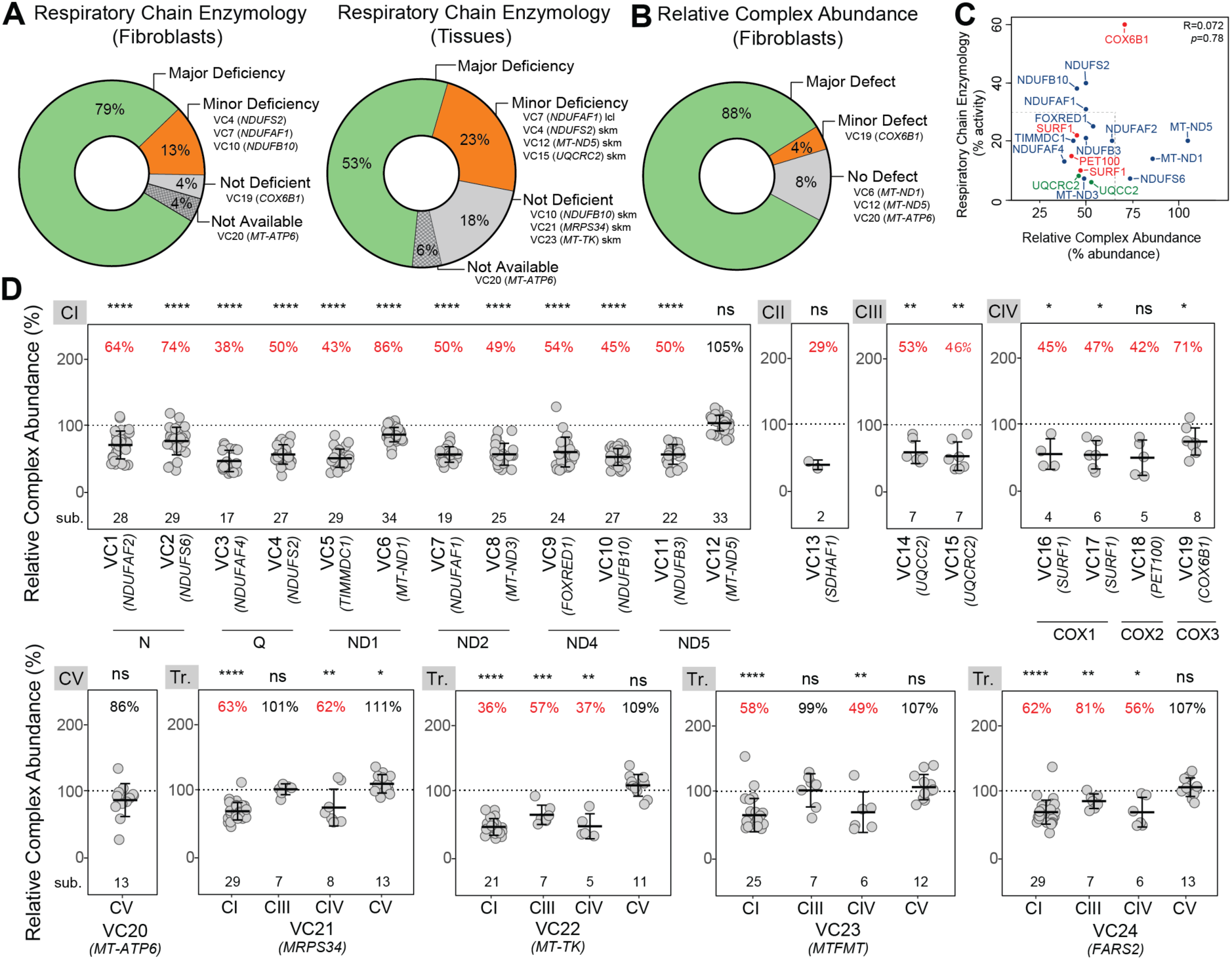
Proteomics outperforms clinical respiratory chain enzymology in detection of mitochondrial disorders. **A –** (left) Summary of the RCE results in fibroblasts according to the Bernier criteria^31^. (right) Summary of the RCE results in tissues (skeletal muscle, skm or lymphoblastoid cell line, LCL) according to the Bernier criteria^31^. **B –** Summary of the RCA results from quantitative proteomic data. **C –** Pearson correlation between RCA results and Respiratory Chain Enzymology results for Complexes I, III and IV. **D –** RCA results of OXPHOS complexes from the validation cohort showing the predicted affected complexes for each cell line. CI-V = Complex I-V. Tr. = translation. Middle bar represents mean complex abundance. Upper and lower bars represent 95% confidence interval. Significance was calculated from a paired t-test between the individual protein means. **** = p<0.0001, *** = p<0.001, ** = p<0.01, * = p<0.05, ns = not significant, p>0.05.

Relative complex abundance (RCA) analysis was performed incorporating the correction for mitochondrial content (**Supplementary Fig. 4**) as described above. We classified a major defect as an RCA abundance ≤65% relative to controls or ≤75% with absent detection of the protein of interest in the proband with >2 peptides detected in controls while minor defects were classified by an RCA abundance of ≤75% relative to controls (**Supplementary Table 2**). Using these criteria, a major RCA defect was detected in 83% of the cell lines in the validation cohort, a minor defect in a single case, VC19 (*COX6B1*), and no defect was detected in three cell lines with mtDNA variants (**Fig. 2B**). Overall, there is no strong correlation between RCE activity and an RCA value relative to control (R=0.072, *p*=0.78) (**Fig. 2C**).

For isolated Complex I disorders, 83% (10/12 patients) had a complex I abundance lower than 75% relative to controls analysed (*p*<0.0001). Two of the three Complex I-deficient fibroblast lines harbouring missense mtDNA-encoded variants were refractory to RCA analysis but diagnostic by RCE. mtDNA variant heteroplasmy in fibroblasts was 92% for VC6 *MT-ND1* and 71% for VC12 *MT-ND5*, highlighting a limitation of our approach in the detection of some mtDNA-driven disorders. Importantly, in all 10 fibroblast lines where RCA was diagnostic for a Complex I disorder, lower abundance (<65% relative to controls) of Complexes III, IV and V were not noted (**Supplementary Fig. 4**). While Complex II was reduced to <75% in four of these, RCE similarly identified a probable Complex II defect for one (*FOXRED1*), supporting the accuracy and specificity of RCA analysis. In 7/7 individuals with isolated Complex II, III or IV disorders, the abundance of the relevant complex was reduced to <75% controls (**Fig. 2D**) however t-test significance varied, especially in CII and IV defects where the number of detected subunits was as low as two. All were specific for the relevant isolated defect except for *UQCRC2* where a Complex I abundance of 67% control (*p*<0.0001) was noted, as expected for Complex III disorders^32^.

For isolated disorders involving Complexes I-IV due to nuclear defects, we determine an RCA value in fibroblasts of <65% controls to be diagnostic. The same threshold may also be applied to isolated disorders caused by pathogenic variants in mtDNA including those leading to Complex V deficiency, however smaller reductions may not exclude pathogenicity as seen in VC6 (*MT-ND1*), VC12 (*MT-ND5*) and VC20 (*MT-ATP6*) (**Fig. 2D**). Interestingly, in the case of VC6 this was likely not due to heteroplasmy, which was 92% NC_012920.1(*MT-ND1*):m.3949C>T in fibroblasts. On the other hand, in VC12 the NC_012920.1(*MT-ND5*):m.13513G>A variant found at 71% in fibroblasts, did not show the expected RCA defect while it satisfied the major criteria in RCE (**Supplementary Table 2**), suggesting impaired enzyme kinetics in the absence of a structural Complex I defect. Finally, we found that defects directly impacting mitochondrial translation such as *MRPS34* fibroblasts presenting with a destabilised mitochondrial ribosome small subunit^25^, *MT-TK* which encodes the mitochondrial lysine transfer RNA^33^*, MTFMT* which encodes the mitochondrial methionyl-tRNA formyltransferase^34^, and *FARS2* encoding mitochondrial phenylalanyl-tRNA synthetase, led to combined Complex I and IV defects that each met criteria of an RCA value <65% control (**Fig. 2D**), consistent with the major criterion observed in RCE (**Supplementary Table 2**). Taken together, fibroblast RCA detected an OXPHOS defect at <65% relative to controls in the expected complex(es) for 20 of 24 fibroblast lines tested (83%) (**Fig. 2B**). One fibroblast line, VC19 (*COX6B1*), had an RCA of 71% for Complex IV relative to controls, which is in line with a previous report showing remaining assembled complex^35^ and the peripheral position of the late assembled COX6B1 subunit in the structure of Complex IV^36, 37^. Three fibroblast lines where a definitive defect was not observed by RCA harboured variants in mtDNA, suggesting that this class of variant may be more refractory to RCA analysis. Despite this, proteomics-based RCA outperformed RCE in the detection of primary mitochondrial disease. It is important to note here that the nature of proteomics as an untargeted approach means that additional analyses can be performed from the same proteomic data, such as reduced abundance of specific proteins encoded by the gene of interest (**Fig. 1E, F**) and co-dependent proteins in structural modules (**Supplementary Fig. 1B**). This is exemplified in the VC2 (*NDUFS6*) case where despite showing a Complex I abundance of 74%, topographical mapping of Complex I subunit abundances against the cryo-EM structure show specific reduction of the N-module of Complex I, providing strong evidence in supporting disease causation.

After benchmarking quantitative proteomics and RCA analysis against the validation cohort, we applied our label-free DIA method to six undiagnosed cases where primary fibroblast or skeletal muscle were available for proteomic analysis.

### UDP1 (*MT-ATP6*)

UDP1 (*MT-ATP6*) was a four-year-old child who had intrauterine growth retardation, truncal hypotonia, microcephaly, global developmental delay, left ventricular noncompaction and Wolff-Parkinson-White syndrome. Biochemically there was a marginal elevation of blood lactate (2.8 mmol/L; normal range 0.7 – 2.0) and 3-methylglutaconic aciduria. Missense variants were identified in *MT-ATP6* [NC_012920.1(*MT-ATP6*):m.8672T>C;p.(Leu49Pro)], a *de novo* variant with 81% heteroplasmy in blood, and *ATAD3A* (heterozygous c.683C>T;p.Thr228Met) from WES plus mtDNA sequencing from blood and both classified as a VUS. Clinical RCE assays typically do not include Complex V activity and proteomics was sought to provide functional evidence. Despite proteomic RCA analysis showing no reduction in abundance of Complex V (104% relative to controls), there was a reduction in the number of Complex V subunits identified from 13 to 10 compared to the diagnosed *MT-ATP6* patient as well as other cases in the same batch (**Fig. 3A** and **Supplementary Fig. 4**). The Complex V subunits that are not detected in UDP1 (*MT-ATP6*) but are detected in VC20 (*MT-ATP6*) are ATP5IF1, the ATPase inhibitor, and ATP5MD (ATP5MK/DAPIT) and ATP5MPL (ATP5MJ/MP68/6.8PL), which like ATP6, are incorporated at a late stage in the assembly of Complex V^36, 38^. The latter suggests a specific turnover of late stage assembled subunits of Complex V below the limit of detection of proteomics in the proband, which contributes to the unchanged Complex V levels seen in the RCA of UDP1 (*MT-ATP6*). We processed the same data using q-value sparse run-wise imputation in the Spectronaut^®^ that allows for peptides that were identified in one sample to be imputed at the limit of detection in samples where they were not detected. This allowed us to estimate the fold changes of ATP6 (*MT-ATP6*) and ATP5MPL proteins in UDP1 (*MT-ATP6*) and correlate their reduced levels with VC20 (*MT-ATP6*) as well as confirming unchanged levels of ATAD3A (**Fig. 3B**). Another data analysis strategy we explored for UDP1 (*MT-ATP6*) was the use of an *in-silico* library-free analysis including the protein sequence of the ATAD3A variant p.Thr228Met. As expected, the peptide containing the methionine at position 228 was only detected in the patient (**Fig. 3C**) and present at ∼43% of the abundance relative to the canonical peptide expressed from the other allele (**Supplementary Fig. 5A**). Taken together, these results suggest that the *MT-ATP6* p.Leu49Pro variant is the likely diagnosis for this case.

**Figure 3.**
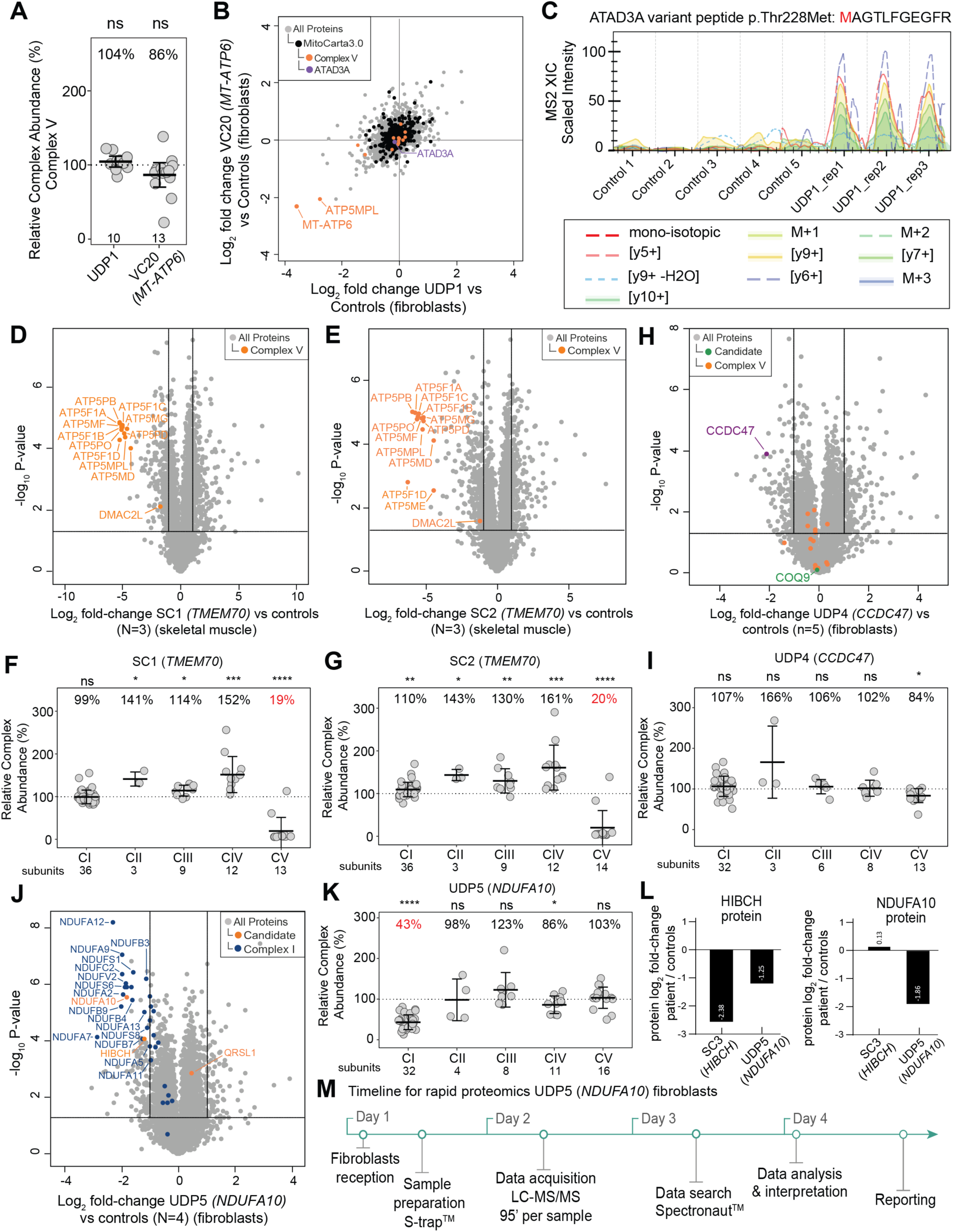
Proteomics supports the diagnosis of patients with suspected mitochondrial disorders. **A –** Relative Complex Abundance (RCA) of Complex V subunits in undiagnosed patient UDP1 (*MT-ATP6*) and diagnosed VC20 (*MT-ATP6*) patient. Middle bar represents mean complex abundance. Upper and lower bars represent 95% confidence interval. Significance was calculated from a paired t-test between the individual protein means. ns = not significant, *p*>0.05. **B –** Correlation between log_2_ fold-changes from whole-cell proteins from run-wise imputed data between undiagnosed patient UDP1 (*MT-ATP6*) and diagnosed patient VC20 (*MT-ATP6*) harbouring mutations in *MT-ATP6* and controls showing reduced abundance in late assembly proteins, MT-ATP6 and ATP5MPL, for both patients. **C –** Spectral intensity of the peptide containing the ATAD3A p.Thr228Met variant detected in the UDP1 patient but not in controls. **D –** Volcano plot of whole cell proteins from SC1 (*TMEM70*) whole-cell skeletal muscle compared to controls (N=3) showing reduced abundance of Complex V subunits (orange dots). Vertical lines represent ± 2-fold-change equivalent and horizontal lines represent significance p-value :: 0.05 equivalent from a two-sample t-test. **E –** Volcano plot of whole cell proteins from SC2 (*TMEM70*) whole-cell skeletal muscle compared to controls (N=3) showing reduced abundance of Complex V subunits (orange dots). Vertical lines represent ± 2-fold-change equivalent and horizontal lines represent significance p-value :: 0.05 equivalent from a two-sample t-test. **F –** Relative Complex Abundance (RCA) of OXPHOS subunits in undiagnosed patient SC1 (*TMEM70*). Middle bar represents mean complex abundance. Upper and lower bars represent 95% confidence interval. Significance was calculated from a paired t-test between the individual protein means. **** = p<0.0001, *** = p<0.001, * = p<0.05, ns = not significant, *p*>0.05. **G –** Relative Complex Abundance (RCA) of OXPHOS subunits in undiagnosed patient SC2 (*TMEM70*). Middle bar represents mean complex abundance. Upper and lower bars represent 95% confidence interval. Significance was calculated from a paired t-test between the individual protein means. **** = p<0.0001, *** = p<0.001, ** = p<0.01, * = p<0.05. **H –** Volcano plot of whole cell proteins from UDP4 (*CCDC47*) fibroblasts compared to controls showing reduced abundance of CCDC47. No significant changes to Complex V proteins (orange). Vertical lines represent ± 2-fold-change equivalent and horizontal lines represent significance p-value :: 0.05 equivalent from a two-sample t-test. **I –** Relative Complex Abundance (RCA) of OXPHOS complexes from UDP4 (*CCDC47*) fibroblasts compared to controls. * = p<0.05, ns = not significant, *p*>0.05. **J –** Volcano plot of whole cell proteins from UDP5 (*NDUFA10*) fibroblasts compared to controls showing reduced abundance of NDUFA10 and other structural subunits of Complex I. Vertical lines represent ± 2-fold-change equivalent and horizontal lines represent significance p-value :: 0.05 equivalent. Blue = Complex I subunits. **K –** Relative Complex Abundance (RCA) of OXPHOS complexes from UDP5 (*NDUFA10*) fibroblasts compared to controls (N=4) showing isolated Complex I defect. Middle bar represents mean complex abundance. Upper and lower bars represent 95% confidence interval. Significance was calculated from a paired t-test between the individual protein means. **** = p<0.0001, * = p<0.05, ns = not significant, *p*>0.05. **L –** NDUFA10 and HIBCH protein abundance from two-sample t-test in SC3, a confirmed *HIBCH* proband and UDP5 (*NDUFA10*) whole-cell fibroblasts compared to controls showing an approximate half reduction in HIBCH levels in UDP5 (*NDUFA10, HIBCH* carrier) compared to SC3 and unchanged NDUFA10 levels in SC3. **M –** Timeline for the rapid proteomics from live fibroblast sample receipt to results for UDP5 (*NDUFA10*) case achieved in less than five days.

The lack of a Complex V defect in proteomics RCA analysis of both *MT-ATP6* cases (VC20 and UDP1) prompted us to analyse additional patients with known pathogenic variants affecting Complex V. Supporting cohort probands SC1 and SC2 were two unrelated infants presenting with lactic acidosis, cardiac abnormalities (persistent patent ductus arteriosus, ventriculo-septal defect and cardiomyopathy), mild dysmorphic facial features, hypospadias, hyperammonaemia and 3-methyl-glutaconic aciduria. Targeted gene sequencing based on this phenotype identified in both individuals a homozygous intronic founder variant NM_017866.6(*TMEM70*):c.317-2A>G classified as Class 5/pathogenic, with several patients previously reported^39–43^. Skeletal muscle samples were available for both cases and compared against three unrelated individuals using the library-free DIA approach. Whole muscle proteomic analyses showed reduction of several Complex V subunits in SC1 and SC2 respectively (**Fig. 3D** and **E**). RCA analyses quantified the relative abundance of Complex V at 19% and 20% (**Fig. 3F and G**), demonstrating that defects in Complex V can be detected using RCA analysis. TMEM70 has also been implicated in Complex I assembly^44, 45^, although no defects in Complex I abundance were noted in these muscle samples.

### UDP4 (*CCDC47*)

UDP4 (*CCDC47*) was born to consanguineous parents, presenting with intrauterine growth restriction, severe failure to thrive, short stature, hypotonia, marked developmental delay, mildly dysmorphic features, severe pruritus woolly hair and increased plasma bile acids (224 μmol/L; normal range <7), Ricketts and decreased mitochondrial CIII activity in liver and muscle^46^. WES performed on DNA extracted from fibroblasts and analysed using a mitochondrial disease gene panel identified single heterozygous missense variants NM_001358921.2(*COQ2*):c.460G>C; p.(Gly154Arg) and NM_020312.4(*COQ9*):c.826C>T p.(Arg276Trp). Suspecting a compound oligogenic mechanism impacting Coenzyme Q biosynthesis, RCA analysis was performed on fibroblast cells. No abundance changes to COQ9 or respiratory chain complexes I-IV were identified, although an abundance of 84% in Complex V was noted (**Fig. 3H** and **I**). This reduction was regarded as not biologically significant as no rare variants in Complex V related genes were flagged from sequencing data. Transcriptomic analysis performed in RNA extracted from skeletal muscle showed a reduced abundance of *CCDC47* transcript **(Supplementary Fig. 5B)**. Re-analysis of proteomics data was sought to provide functional evidence to support the transcriptomic findings, which confirmed a reduction in the abundance of CCDC47 in primary fibroblasts from imputed data (**Fig. 3H**). Re-analysis of sequencing data identified a homozygous variant NM_020198.3(*CCDC47*):c.431C>G;p.(Thr144Asn). SpliceAI predicted (δ score 0.964) the activation of a donor-splice site five nucleotides upstream of this variant, and PCR analysis of the muscle cDNA showed the presence of both the missense transcript and a second shorter one, missing 121 bp from the 3’ end of exon 4 resulting in a loss-of-function type transcript p.(Val143Glyfs*15) **(Supplementary Fig. 5C)**. This variant is not observed in the gnomAD population database nor any clinical cases previously but is predicted to be classified as likely pathogenic/Class 4, supported by the observed splicing defect, as well as reduced abundance in *CCDC47* transcript and CCDC47 protein levels. CCDC47 has been previously linked to Trichohepatoneurodevelopmental Syndrome^47^ (MIM 618268) which presents an overlapping phenotype to UDP4 (*CCDC47*). The observed reduction in *CCDC47* transcripts due to incorrect splicing is in keeping with previously reported patients, all of whom harbour loss-of-function variants^47, 48^.

### UDP5 (*NDUFA10*)

UDP5 presented with nystagmus, hypotonia, vomiting and weight loss at 20 weeks of age. She had lactic acidosis, concentric left ventricular hypertrophy with 48% ejection fraction, T2 hyperintense lesions in the substantia nigra and passed away at 24 weeks of age. Targeted exome sequencing analysis flagged candidate variants NM_004544.4(*NDUFA10*):c.914T>C; p.(Leu305Pro) homozygous VUS/Class 3, NM_014362.4(*HIBCH*):c.891+1G>A, heterozygous likely pathogenic/Class 4 and NM_018292.5(*QRSL1*):c.22G>C, p.(Glu8Gln) heterozygous VUS/Class 3. Fibroblast proteomics detected NDUFA10 and HIBCH below 2-fold-change (representing >50% reduction) in the patient relative to controls (**Fig. 3J and Supplementary Figure 5D**). QRSL1 level was slightly increased but not significant, which led us to further investigate *NDUFA10* and *HIBCH* leads. NDUFA10 is a structural subunit of Complex I and additional functional evidence for pathogenicity could be drawn from reduced abundance of other Complex I subunits, with RCA analysis quantifying Complex I abundance at 43% in UDP5 (*NDUFA10*) compared to controls, meeting the criteria for a major defect (**Fig. 3K**). We further assessed whether the levels of HIBCH protein were compatible with a carrier (heterozygous) state by analyzing fibroblasts from an unrelated patient (SC3) diagnosed with bi-allelic pathogenic variants: NM_014362.4(*HIBCH*):c.891+1G>A & NM_014362.4(*HIBCH*):c.470G>A; p.(Arg157Gln). Proteomic results show that HIBCH protein in SC3 (*HIBCH*) is almost two times less abundant than in UDP5 (*NDUFA10*) (**Fig. 3L, left panel**) while the levels of NDUFA10 protein were unchanged in SC3 (*HIBCH*), suggesting that a reduction in NDUFA10 is not a secondary defect from a primary *HIBCH* defect (**Fig. 3L right panel**). The lack of any decrease in Complex I in SC3 (*HIBCH*) suggests that the phenotype in UDP5 (*NDUFA10*) is likely to have arisen from the *NDUFA10* variant while the *HIBCH* variant is expressed as a carrier and further investigation for a second missed variant in *HIBCH* was not undertaken. UDP5 was the first child of non-consanguineous parents of Māori ethnicity. The mother was 6 weeks pregnant with a second child at the time of analysis and the family sought prenatal genetic testing (PGT). With this in mind, we prioritised sample processing and analysis, and were able to return results in less than 4 days from receipt of fibroblasts (**Fig. 3M**) with the results sufficient to support the upgrade of the *NDUFA10* variant c.914T>C; p.(Leu305Pro) to likely pathogenic/Class 4. This facilitated PGT, which confirmed absence of the *NDUFA10* variant, allowing continuation of the pregnancy and delivery of a healthy child who, at the time of writing, is 15-months old.

The rapid turnaround time (TAT) in this case was only possible as fibroblasts from UDP5 were available for analysis at the time of proteomics testing. Establishing and culturing a new fibroblast line typically takes up to ∼2 months and requires a skin biopsy^49^ which is increasingly considered invasive and can potentially delay a diagnosis. We sought to explore the feasibility of rapid proteomics-based testing for suspected mitochondrial disorders using peripheral blood mononuclear cells (PBMCs), which can be readily obtained from whole blood and have been used in various diagnostic functional testing approaches previously. To test the applicability of PBMCs in proteomic testing we first generated a fractionated spectral library and compared it against the fibroblast spectral library and coverage of Mendeliome proteins (**Fig. 4A**). The PBMC library was generated from peptides of 39 normal individuals aged 0-17 years of age and can be used to confidently identify 8,016 gene products, including 52% of known Mendeliome genes^27^. In comparison, our fibroblast spectral library covers 8,305 genes and 55% of the Mendeliome. We performed a Principal Component Analysis (PCA) of whole-cell proteins in the 37 PBMC samples showing that over 99% of the proteins are tightly clustered at the whole-cell level and over 97% at the mitochondrial level (**Fig. 4B**).

**Figure 4.**
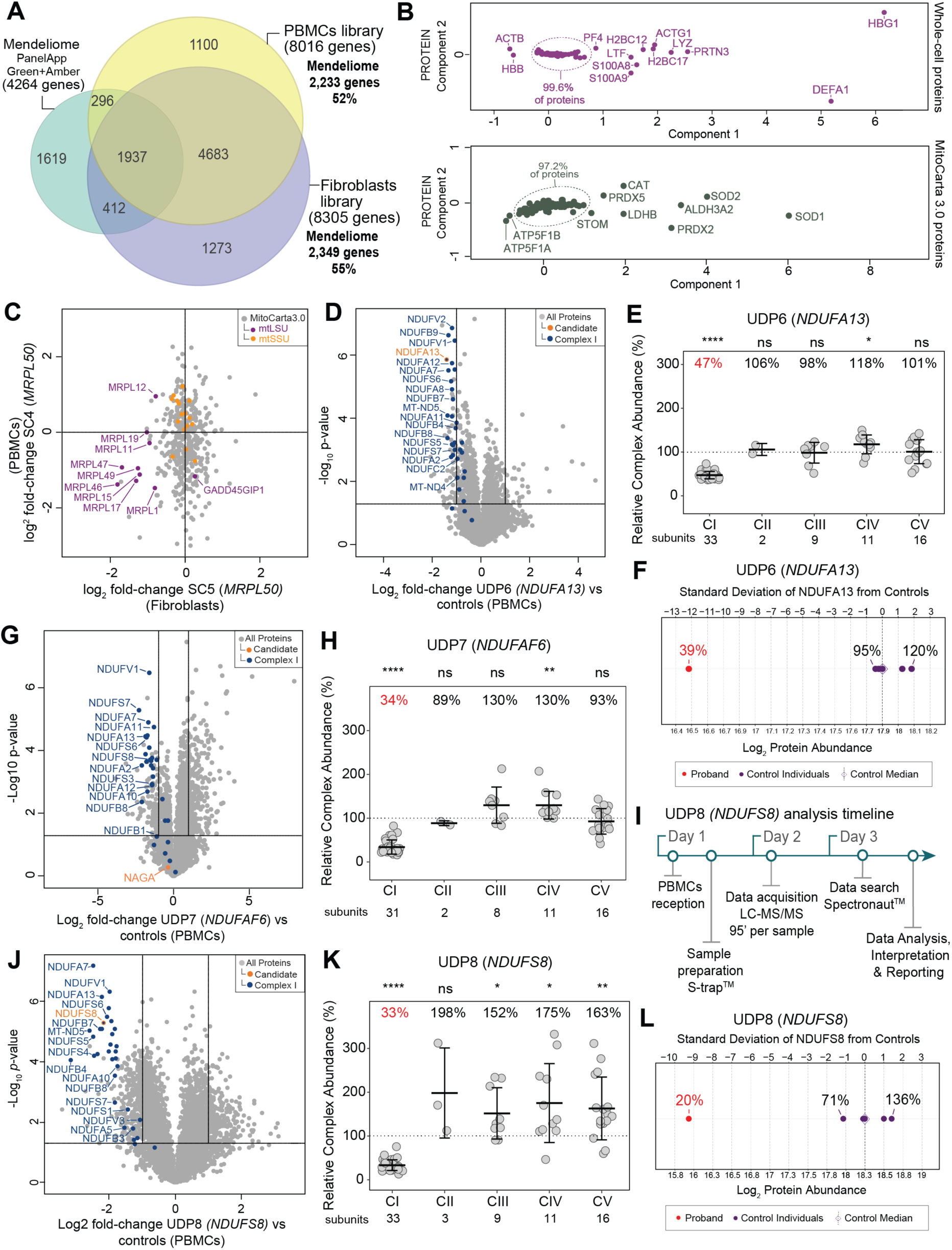
Utility of ultra-rapid proteomics supports the diagnosis of critically ill infants with suspected mitochondrial disorders. **A –** Venn diagram showing the coverage of Mendeliome genes (PanelApp Australia Green and Amber entries, 4,264 genes) in the fibroblast spectral library (55% of Mendeliome list of genes) and PBMC library (52% of Mendeliome list of genes). **B –** Principal Component Analysis (PCA) of the pilot PBMC normative data (N=36) for whole-cell proteins (top panel) and MitoCarta3.0 proteins (lower panel). **C –** Scatter plot showing the correlation between PBMC (SC4) and fibroblast (SC5) samples based on log_2_ fold-changes from whole-cell proteins in a diagnosed patient with variants in *MRPL50* relative to controls^21^ showing reduced abundance of the proteins belonging to the large subunits of the mitochondrial ribosome (mtLSU; purple), mtSSU = mitoribosome small subunit (orange). **D –** Volcano plot of whole cell proteins from UDP6 (*NDUFA13*) PBMCs compared to controls (N=5) showing reduced abundance of NDUFA13 (orange dot) and other structural subunits of Complex I. Vertical lines represent ± 2-fold-change equivalent and horizontal lines represent significance p-value :: 0.05 equivalent. Blue = Complex I subunits**. E –** Relative Complex Abundance (RCA) of OXPHOS complexes from UDP6 (*NDUFA13*) PBMCs compared to controls (N=5) showing an isolated Complex I defect. Middle bar represents mean complex abundance. Upper and lower bars represent 95% confidence interval. Significance was calculated from a paired t-test between the individual protein means. **** = p<0.0001, * = p<0.05, ns = not significant, *p*>0.05. **F –** Protein range for NDUFA13 in PBMCs in UDP6 (*NDUFA13,* red dot) and controls (N=5, purple dots) showing standard deviation of -12.2 from the control median**. G –** Volcano plot of whole cell proteins from UDP7 (*NDUFAF6*) PBMCs compared to controls (N=5) showing reduced abundance of NDUFS8 and other structural subunits of Complex I. Vertical lines represent ± 2-fold-change equivalent and horizontal lines represent significance p-value :: 0.05 equivalent. Blue = Complex I subunits. **H –** Relative Complex Abundance (RCA) of OXPHOS complexes from UDP7 (*NDUFAF6*) PBMCs compared to controls (N=5) showing an isolated Complex I defect. Middle bar represents mean complex abundance. Upper and lower bars represent 95% confidence interval. Significance was calculated from a paired t-test between the individual protein means. **** = p<0.0001, ** = p<0.01, ns = not significant, *p*>0.05. **I –** Timeline for ultra-rapid proteomics from PBMC sample receipt to results for UDP8 (*NDUFS8*) case achieved in less than 3 business days. **J –** Volcano plot of whole cell proteins from UDP7 (*NDUFS8*) PBMCs compared to controls (N=5) showing reduced abundance of NDUFS8 protein (orange dot) and other structural subunits of Complex I. Vertical lines represent ±2-fold-change equivalent and horizontal lines represent significance p-value :: 0.05 equivalent. Blue = Complex I subunits**. K –** Relative Complex Abundance (RCA) of OXPHOS complexes from UDP8 (*NDUFS8*) PBMCs compared to controls (N=5) showing isolated Complex I defect. Middle bar represents mean complex abundance. Upper and lower bars represent 95% confidence interval. Significance was calculated from a paired t-test between the individual protein means. **** = p<0.0001, ** = p<0.01, * = p<0.05, ns = not significant, *p*>0.05. **L –** Protein range for NDUFS8 in PBMCs in UDP8 (*NDUFS8,* red dot) and controls (N=5, purple dots) showing standard deviation of -9.3 from the control median.

Next, we validated the use of PBMCs against fibroblasts by analysing both sample types from a diagnosed patient with pathogenic variants in *MRPL50* (Twin 2 in ^21^, SC4 and SC5). MRPL50 is a structural subunit of the mitoribosome^50^ and defects in this subunit lead to impaired assembly of the large mitoribosomal subunit (mtLSU)^21^. We performed a correlation of the log_2_ fold-change levels of mitochondrial proteins identified in both samples relative to controls which showed reduced abundance of multiple proteins belonging to the mtLSU in both sample types (**Fig. 4C**). RCA quantified the abundance of the mtLSU in 63% and 48% in PBMCs and fibroblasts respectively **(Supplementary Fig. 5E**).

### UDP6 (*NDUFA13*)

UDP6 (*NDUFA13*) presented at 2 years of age with chronic ataxia. She was the first child to non-consanguineous healthy parents from Iran and had a healthy 2-month-old brother. There was no family history of any neurological issues. She presented with chronic, stable ataxia, having been unsteady and falling frequently since she started walking at 14 months of age. She had mild fine motor delay and a tremor. Her social, language, and cognitive development was age appropriate. On examination, UDP6 (*NDUFA13*) had normal growth parameters and was not dysmorphic. She had mild gait ataxia with a broad base. Her tone, power and reflexes were normal in upper and lower limbs bilaterally.

Ophthalmology examination was unremarkable. Magnetic resonance imaging (MRI) of brain showed bilateral symmetrical T2/FLAIR hyperintensity of the cerebral peduncles of the midbrain (and possibly the substantia nigra) and two symmetrical foci of high signal in the dorsal medulla with diffusion-weighted imaging (DWI) restriction. There were no basal ganglia or thalamic abnormalities. This was suggestive of a mitochondrial disorder. Serum blood lactate was elevated 2.4 (RR 1.0-1.8). Other metabolic investigations were normal. Audiology assessments and echocardiogram were normal. At 4 years of age, she has ongoing mild ataxia that improved with physiotherapy and was no longer falling. She has recently had deterioration in her vision and been diagnosed with optic neuropathy. Trio exome sequencing identified compound heterozygous variants NM_015965.7(*NDUFA13*):c.170G>A; p.(Arg57His) likely pathogenic/Class 4 and NM_015965.7(*NDUFA13*):c.187G>A; p.(Glu63Lys) VUS/Class 3. Proteomics was sought to provide functional evidence for the VUS/Class 3 variant and blood was collected for PBMC isolation as fibroblasts were not available at the time of analysis. Five unrelated PBMCs from age-matched normal donors were used as controls. A volcano plot showed reduction of NDUFA13 protein and several other Complex I subunits in UDP6 (*NDUFA13*) (**Fig. 4D**) and RCA quantified the isolated Complex I defect at 47% control satisfying the criteria for a major defect (**Fig. 4E**). Analysis of the level of the NDUFA13 protein in the proband demonstrated a residual abundance of 39%, which corresponds to greater than 12 standard deviations below the control median (**Fig. 4F**). Proteomics data were subsequently used as functional evidence to upgrade the NM_015965.7(*NDUFA13*):c.187G>A;p.(Glu63Lys) variant to likely pathogenic/Class 4.

### UDP7 (*NDUFAF6*)

UDP7 (*NDUFAF6*) was referred to the study at five months of age with asymmetric early onset growth restriction, central hypotonia, proximal renal tubular dysfunction, macrocytic anaemia, severe exocrine pancreatic insufficiency, liver dysfunction with cholestasis and mild persistent elevated lactate in CSF and blood. MRI showed symmetrical diffusion abnormality involving corticospinal tract, areas of brainstem and medial cerebellar hemisphere. He died at seven months of age due to progressive bulbar dysfunction with a probable clinical diagnosis of Leigh syndrome. Clinical trio WES identified a single pathogenic (Class 5) heterozygous NM_000262.3(*NAGA*):c.973G>A; p.(Glu325Lys) variant, and two *in trans* variants in *NDUFAF6*, a frameshift NM_152416.4(*NDUFAF6*):c.267delT, p.(Tyr323Ilefs*18) variant and a deep intronic variant, NM_152416.4(*NDUFAF6*):c.298-768T>C, both classified as VUS/Class 3. PBMCs were the sample of choice due to lower invasiveness and potential for fast turnaround. Whole-cell proteomics identified no changes in the abundance of NAGA protein relative to controls (**Fig 4G**). *NAGA* encodes the alpha-N-acetylgalactosaminidase, a lysosomal enzyme whose deficiency causes a rare autosomal recessive lysosomal storage disorder. Previous reports of NAGA-deficiency show heterogenous presentations ranging from infantile-onset neuroaxonal dystrophy (Schindler disease type I, MIM <u>609241</u>) to an adult-onset disorder characterised by angiokeratoma corporis diffusum and mild intellectual impairment (Schindler disease type II, also known as Kanzaki disease MIM <u>609242</u>)^51^. In addition to the proteomic studies, UDP7 *(NDUFAF6)* did not appear to have clinical features consistent with NAGA-deficiency, and infants with this condition typically develop normally until about a year old. We then turned our investigations to *NDUFAF6* (*C8orf38*) which encodes an assembly factor involved in biogenesis of the core peripheral arm subunit NDUFS8 of Complex I^52^ and has previously been associated with the stability of the ND1 module of Complex I^53,54^. Previous reported cases of pathogenic variants in *NDUFAF6* present with a range of symptoms, including Leigh syndrome and the Acadian variant of Fanconi syndrome 5 (proximal renal tubular dysfunction) (FRTS5, MIM 618913)^53–57^. UDP7 *(NDUFAF6)* had clinical features consistent with Leigh syndrome, and while the aetiology of his liver dysfunction and severe exocrine pancreatic insufficiency was somewhat less clear, both would broadly fit with a mitochondrial cytopathy noting Complex I deficiencies often do not correlate perfectly with a single clinical subdivision. The NM_152416.4(*NDUFAF6*):c.298-768T>C variant has been reported as likely pathogenic previously, specifically in relation to the FRTS5 phenotype^56^. A volcano plot from whole-cell PBMC proteomics data showed reduction in the abundance of several subunits of Complex I in UDP7 (*NDUFAF6*), including NDUFS8, (**Fig. 4G**) with RCA analysis quantifying the isolated Complex I defect with a major defect of 34% relative to controls (**Fig. 4H**). Functional proteomic data and phenotype match support *NDUFAF6* as being the causative gene for this clinical case, and the VUS/Class 3 variants are in the process of re-curation at the time of writing.

### UDP8 (*NDUFS8*)

UDP8 (*NDUFS8*) was a neonate admitted to hospital presenting with persistent lactic acidemia, hypertrophic cardiomyopathy, left pulmonary artery stenosis, thickened pulmonary valve, hypotonia and microcephaly with progressive deterioration from 4 weeks of age, succumbing at 5 weeks of age. Brain MRI showed abnormalities in the corpus callosum and diffuse increased signal throughout the supratentorial white matter with small foci of gliosis in the peritrigonal regions. Ultra-rapid trio WGS identified a homozygous intronic variant NM_002496.4(*NDUFS8*):c.501+5G>A with biparental inheritance. Due to the critical deteriorating condition, blood was collected from the proband and subjected to ultra-rapid proteomics. Five unrelated age-matched controls were used for comparison and the complete analysis was performed with a 54-hour turnaround time (TAT) from PBMC sample reception to the reporting of results to the clinical team (**Fig. 4I**). Whole-cell proteomic analysis of PBMCs showed significant reduction of several Complex I subunits, including NDUFS8 (**Fig. 4J**) and RCA analysis showed an isolated Complex I defect with a major defect of 22% relative to controls (**Fig. 4K**). Residual abundance of the NDUFS8 protein in the proband was quantified at 20%, representing over 9 standard deviations below the control median (**Fig. 4L**). The strong phenotype match together with functional evidence from proteomics support disease causation due to variants in *NDUFS8* and leading to variant upgrade to likely pathogenic/Class 4.

## Discussion

The estimated prevalence of rare disease in the population is in the range of 3.5-6%, equivalent to 263-446 million individuals worldwide^58^. There are over 7,000 different rare diseases known, with a current diagnostic yield of ∼35-70% from WES or WGS^1–5^. Mitochondrial disease is a group of rare diseases caused by variants in over 300 known genes^6, 16^, where RCE has been historically performed to confirm a specific OXPHOS defect in the functional validation of variant pathogenicity^16^.

We have previously used an approach we term relative complex abundance (RCA) where proteomics data are used to quantify the residual abundance of OXPHOS complexes in clinical samples from individuals suspected of mitochondrial disease. We have shown that RCA acts as an effective proxy for clinically accredited RCE, including when appropriate samples have not been available^20–22, 24, 25, 59–61^. In an RCA analysis, the abundance of a complex in a sample is calculated from the mean of each individual subunit abundance detected by more than two peptides across each complex. The power of RCA analysis relies on the quantification of multiple peptides per protein in a complex with a high degree of subunit stability co-dependence, as we and others have previously shown for Complex I, III, IV, the mitoribosome^11, 17, 20–22, 25, 59, 60, 62, 63^ and non-mitochondrial complexes such as the exocyst and nuclear pore complexes^64, 65^. RCE, on the other hand, typically relies on enzyme rate estimates determined in several replicates of patient samples and compared against age-matched controls. In some instances, we have found RCA demonstrated higher sensitivity by detecting a specific OXPHOS defect where RCE was inconclusive^20, 22^. RCA can also show higher specificity for use in functional validation of variant pathogenicity by detecting reduction of specific parts of the complex (e.g. loss of subunits in the catalytic Complex I N-module in VC1-*NDUFAF2* and VC2-*NDUFS6*), implicating only the 10 N-module subunits as candidate genes compared with loss of Complex I RCE potentially being caused by variants in any of more than 50 genes encoding Complex I subunits and assembly factors^26, 66^. In other cases, some variants encoded by the mtDNA resulted in no defect on RCA analysis despite over 70% and 90% heteroplasmy levels in fibroblasts. This suggests that some mtDNA variants can be refractive to RCA analysis due to an enzymatic and not a structural defect, where RCE might be an appropriate test to provide functional evidence. In terms of variant type, mitochondrial DNA variants account for only 0.54% of rare diseases while autosomal recessive variants account for over 41%^67^ and are more likely to result in a loss-of-function protein^68^, which can be detected via quantitative proteomics.

RCE is typically performed on a tissue biopsy sample (e.g. skeletal muscle, liver) or primary cultured cells (e.g. fibroblasts). Establishing and growing primary fibroblasts from skin biopsies to a sufficient volume of cells for RCE analysis involves weeks to months of cell culture^49^, and, in some cases, tissue specificity results in only mild or undetectable defects as measured by RCE^10^. Here we have demonstrated the feasibility of using PBMCs in diagnostic proteomics testing. In our hands, sufficient PBMCs for these analyses can be readily obtained from as a little as 1 ml of whole blood and the process from PBMC isolation to data acquisition performed in less than 48 hours. Together these properties and the results presented in this study demonstrate that PBMCs can be combined with our ultra-rapid proteomics pipeline for functional validation of variants causing defects in the mitochondrial OXPHOS system and the mitoribosome.

The untargeted nature of quantitative proteomics also allows this technique to be applied beyond mitochondrial disease, as we have shown here for a mitochondrial disease phenocopy (UDP4; *CCDC47*) and elsewhere for nuclear pore, neuronal exocytic vesicle trafficking and rigid spine disorders^64, 65, 69^. Over 50% of known disease-associated genes^27^ are routinely detected by our quantitative proteomics pipeline in PBMCs or fibroblasts, and over 45% of all disease genes listed in OMIM are part of a protein complex^16^. This means that pathogenic variants in one of these genes can lead to reduction of the protein of interest, which in some cases will result in downstream complex reduction as an additional layer of functional evidence to support variant upgrade. In terms of variant type, we show that quantitative proteomics can provide functional evidence for a wide range of genetic variants including CNV^62^, splice site and deep intronic^20, 22^, and missense variants^21^. Missense variants are the most common type of variant accounting for 60% of pathogenic variants associated with autosomal recessive disorders^18^ and are usually refractory to transcriptomic analysis. For some cases, proteomics can also provide supportive functional evidence supportive when the protein encoded by the candidate gene is not detectable, either due to protein abundance being below the limit of detection or for genes encoding tRNAs, as seen in VC22 (*MT-TK*), further expanding the potential application of proteomics in rare disease diagnosis.

A genomics-first approach to rare disease diagnosis has markedly increased diagnostic rates and, in the case of rapid and ultra-rapid genomic testing, shortened diagnostic odysseys to days instead of months^65, 70–72^. In the case of mitochondrial and many other rare diseases, diagnoses are often now achieved using just a blood sample, sparing many patients from an invasive biopsy of muscle or other tissues. As a result, diagnostic centres such as ours now receive a third or less of the number of samples for RCE testing than received 10 years ago. This provides an incentive for untargeted functional tests, such as RNA sequencing and proteomics, that could be financially viable and widely available in a pathology-certified context.

This study also highlights the limitations of quantitative proteomics in detecting abundance changes in variants leading to catalytic defects and some mtDNA-encoded variants in fibroblasts. Moreover, the detection of proteins is related to the limit of detection as determined by the liquid chromatography, mass-spectrometry instrumentation, and data acquisition methods used. However, recent advances in mass-spectrometry instrumentation including development of the asymmetric track lossless (Astral) analyzer offer further improvements in sensitivity and greatly reduced run times^73^. Quantitative proteomics thus offers a further paradigm shift by providing functional evidence for variants in thousands of genes in a single test. This driver will facilitate the translation of proteomics testing for rare diseases into certified pathology laboratories. Clinically delivered proteomics can potentially replace hundreds of tests targeted to specific diseases, which are usually restricted to research settings and carry less weight in the upgrade of variants. Our study also demonstrates that proteomics can be delivered alongside ultra-rapid genomic sequencing approaches to provide functional data in a clinically relevant timeline.

## Materials and Methods

### Ethics statement

This study was conducted in accordance with the revised Declaration of Helsinki and following the Australian National Health and Medical Research Council statement of ethical conduct in research involving humans. Samples were obtained after receiving written, informed consent for diagnostic or research investigations from the respective responsible human ethics institutional review boards. HREC/RCH/34228, HREC/RCH/34183, HREC/89419/RCHM-2022 and HREC/82160/RCHM-2022 were approved by the Royal Children’s Hospital, Melbourne, Ethics in Human Research Committee. HREC/16/MH/251 was approved by the Melbourne Hospital Ethics in Human Research Committee. The REC reference 2002/205 by the Newcastle and North Tyneside Local Research Ethics Committee.

### Genomic investigations

Genomic investigations resulting in diagnosis of patients within the validation cohort (VC) are described in the publications indicated by PMID within **Supplementary Table 1**, with the exceptions of VC13 (*SDHAF1*), VC15 (*UQCRC2*) and VC24 (*FARS2*) which are as follows; Individual VC13 (*SDHAF1)* underwent clinical singleton WES through Baylor Genetics as previously described^74^. For individual VC15 (*UQCRC2*), homozygosity mapping from Illumina HumanCytoSNP-12 v2.1 array data identified substantial long contiguous stretches of homozygosity (LCSH) accounting for ∼3.7% of the genome. Due to the observed complex III RCE defect in this patient, candidate gene sequencing of exons using genomic DNA through PCR and sanger sequencing was performed on complex III-related genes *UQCC1* and *UQCRC2* which lay within these regions of LCSH (primers available upon request). Blood DNA for individual VC24 (*FARS2*) underwent clinical singleton WES testing through the Victorian Clinical Genetics Service (VCGS) as previously described^75^. mtDNAseq was also used to quantify mtDNA variant heteroplasmy levels for individuals VC6 (*MT-ND1*) and VC12 (*MT-ND5*) as previously described^75^. Individuals within the undiagnosed patient (UDP) cohort were investigated as follow: blood DNA from UDP1 (*MT-ATP6*) underwent WES with mitochondrial DNA sequencing (mtDNAseq) as part of the Australian Genomics Mitochondrial Flagship as described^75^. For individual UDP4 (*CCDC47*), sequencing and analysis was performed as previously described^25^. Individual UDP5 (*NDUFA10*) had the Comprehensive Metabolism Panel performed by Blueprint Genetics. Individuals UDP6 (*NDUFA13*) and UDP7 (*NDUFAF6*) both underwent clinical trio WES through the Victorian Clinical Genetics Service (VCGS)^75^. UDP8 (*NDUFS8*) had acute-care rapid trio WGS and analysis at the VCGS as previously published^65^.

### Respiratory chain enzymology

Respiratory chain enzyme activities in fibroblasts and skeletal muscle were measured by spectrophotometry as described^10^. Complex I (CI) was measured as rotenone-sensitive NADH:coenzyme Q_1_ oxidoreductase, Complex II (CII) as succinate:coenzyme Q_1_ oxidoreductase, Complex III (CIII) as decylbenzylquinol:cytochrome *c* reductase, and Complex IV (CIV) as cytochrome *c* oxidase. Citrate synthase (CS) was measured as production of coenzyme A (CoA.SH) from oxaloacetate using the thiol reagent 5,5’-dithio-bis-(2-nitobenzoic acid). Enzyme activities were calculated as initial rates (CI, CII, and CS) or as first-order rate constants (CIII and CIV)^10^.

### Cell Culture Conditions

Fibroblast and HEK293T cell lines were cultured in Dulbecco’s Modified Eagle Medium (DMEM) High Glucose, Sodium Pyruvate and Glutamine (Sigma-Aldrich) supplemented with 10% (v/v) Fetal Calf Serum (FCS; CellSera), 100 U/mL Penicillin-Streptomycin (Gibco) and 50 µg/mL Uridine. Cells were maintained at 37°C with 5% CO2.

### Generation of knockout lines

The NDUFS2^KO^ cell line was generated using a CRISPR/Cas9 plasmid encoding the guide RNA 5’-TGAGGGCTTTGTGCGGCTTCCGG-3’ cloned into the pSpCas9-2A-GFP vector ^26^. The CRISPR was transfected into HEK293T cells and single cells were obtained by Fluorescence-activated cell sorting at the Monash Flow Cytometry Platform (Flowcore). Singe cell populations were screened by loss of cell viability on galactose-containing DMEM and subsequently validated by the loss of protein by western blotting and indel sequencing, which identified two deletions c.[18_36del] and c.[17_42del] Supplementary table 1). The UQCRC2^KO^ cell line was generated using a CRISPR/Cas9 plasmid encoding the guide RNA 5’-CACCGGTACTTACACATCACCCCGC designed using the CHOPCHOP tool and subcloned into pSpCas9(BB)-2A-GFP vector as previously described^26^. The plasmids were validated by sequencing, and then transfected into target cell lines using Lipofectamine™ 3000 Transfection Reagent (Thermo Fischer Scientific) according to manufacturer’s guidelines. Single cells expressing GFP were isolated on a FACS Aria Fusion (BD Biosciences) cell sorter and clonal populations were subsequently screened for relevant gene knockouts using SDS-PAGE and immunoblotting and indel sequencing, which identified two deletions c.[9141_9199del] and [9146_9167del] (Supplementary table 1). The SURF1^KO^ cell line was generated using a CRISPIR/Cas9 system with the HEK293T parental cell line. Two guide RNAs targeting the 5’UTR and exon 2 regions of *SURF1* were cloned into individual pSpCas9-2A-GFP vectors (Addgene, plasmid 48138 ^18^). Both constructs were transfected simultaneously into HEK293T cells using Lipofectamine 3000 (Thermo Fisher Scientific), according to the manufacturer’s instructions. Verification of successful HEK293T SURF1^KO^ was performed via three primer screening PCR were used to amplify incorporated indels via PCR and by SDS-PAGE immunoblotting. The product of external Primers (outside the double CRISPR target sites) underwent M13 primer Sanger Sequencing of alleles, using the Topo^TM^ TA Cloning Kit (Thermo Fisher Scientific) as per the manufacturer’s instructions. Sequencing identified two deletions [c.18_219del] and [c.18_221delinCG] (Supplementary table 1). The ATP5PD^KO^ cell line was generated using a CRISPR/Cas9 plasmid encoding the guide RNA 5’-CACCGCCTTTCCTTGTGGGCAGGT designed using the CHOPCHOP tool and subcloned into pSpCas9(BB)-2A-GFP vector as previously described^26^. The plasmids were validated by sequencing, and then transfected into target cell lines using Lipofectamine™ 3000 Transfection Reagent (Thermo Fischer Scientific) according to manufacturer’s guidelines. Single cells expressing GFP were isolated on a FACS Aria Fusion (BD Biosciences) cell sorter and clonal populations were subsequently screened for relevant gene knockouts using indel sequencing, which identified a single deletion c.[33481insT] (Supplementary table 1).

### PBMC isolation from whole blood

Neonatal blood samples were collected from healthy neonates following routine intramuscular administration of Vitamin K. For older infants and children ages 30 days to 18 years, venous blood samples were obtained from hospital outpatients undergoing minor elective day surgery. An outline of the study protocol and complete detail of participant recruitment, inclusion and exclusion criteria and sample collection are described in our previously published HAPPI Kids study protocol^76^. PBMCs were isolated from fresh blood samples of the 37 HAPPI Kids controls using Ficoll^®^-Paque Plus (GE Healthcare Life Sciences) according to the instructions of the manufacturer with manual removal of the PBMC layer using a Pasteur pipette. For the UDP6 (*NDUFA13*), UDP7 (*NDUFAF6*) and UDP8 (*NDUFS8*) probands, PBMC isolation using Ficoll^®^-Paque Plus was performed in SepMate^TM^ tubes (STEMCELL Technologies), based on manufacturer’s protocols with moderate braking applied for both washes, and a reduction in centrifugation speed and duration to 200 *g* for five minutes for the second wash.

*Data-Independent Acquisition Mass Spectrometry of Fibroblast, HEK293T and PBMCs Cohorts and Data Analysis*

### Spectral Library Sample Preparation

A HEK293T spectral library was generated from both whole cell and mitochondrial HEK293T samples to search data-independent acquisition (DIA) HEK293T knockout cell lines. A total of 100 µg of mitochondrial sample isolated from wildtype HEK293T cells underwent carbonate extraction at pH 11.5 to separate soluble and integral mitochondrial proteins, as previously described^77^. The resulting pellet and supernatant samples, as well as 25 µg whole cell HEK293T sample, were solubilized in lysis buffer for a final composition of 5% SDS and 50 mM Triethylammonium bicarbonate (TEAB) pH 8.5. The lysed whole cell, mitochondrial pellet and supernatant samples were processed using S-trap^TM^ Micro Spin Columns (ProtiFi) according to the manufacturer’s instructions, where samples were reduced with 40 mM 2-chloroacetamide (CAA, Sigma) and alkylated with 10 mM tris(2-carboxyethyl)phosphine hydrochloride (TCEP; BondBreaker, Thermo Fischer Scientific). Isolated proteins were digested at 1:25 trypsin to protein ratio at 37°C overnight and eluted peptides were dried down using a CentriVap Benchtop Vacuum Concentrator (Labconco). Fibroblasts and PBMCs spectral libraries were generating from whole cell samples of either five healthy fibroblast controls or PBMC pellets isolated from 39 healthy individuals ranging from 0-17 years old. Fibroblast or PBMC pellets were solubilized 5% SDS and 50 mM TEAB pH 8.5, and total protein determined by Pierce bicinchoninic acid (BCA) Protein Assay Kit (Thermo Fisher Scientific). A total of 25 µg for each control was processed using the S-Trap^TM^ Micro Spin Columns (ProtiFi), as described above.

Peptides from HEK293T, fibroblast and PBMC samples were reconstituted in 0.5% formic acid (FA) for fractionation. Fibroblast and PBMC control samples were pooled upon reconstitution to create a single fibroblast, HEK293T and PBMC peptide mixture and loaded onto a strong cation exchange (Empore Cation Exchange-SR, Supelco Analytical) stage-tips made as described^78^. Stage tips were washed with 20% acetonitrile (ACN) and 0.5% FA and eluted over seven fractions of increasing concentrations of ammonium acetate (45-300 mM), 20% ACN and 0.5% FA for whole cell fibroblasts, PBMC and HEK293T samples. HEK293T mitochondrial and supernatant samples, were collected over five fractions. All samples underwent a final elution with 5% ammonium hydroxide and 80% ACN, followed by concentration using a CentriVap Benchtop Vacuum Concentrator (Labconoco). Fractions were desalted on SDB-XC (poly(styrene-divinylbenzene); Supelco Analytical) stage tips made in-house as previously described^78^.

### Spectral Library Mass Spectrometry Data Dependent Acquisition

Fractions were reconstituted in 0.1% trifluoracetic acid (TFA) and 2% ACN and each library was analyzed by liquid chromatography (LC)-tandem mass spectrometry (MS/MS) on an Orbitrap Eclipse Mass Spectrometer (Thermo Fischer Scientific) operating on DDA mode over a 125-minute gradient. Tryptic peptides were loaded onto an Acclaim Pepmap nano-trap column (Dinoex-C18, 100 Å, 75 µm × 2 cm) at an isocratic flow of 5 µl/min of 2% ACN and 0.1% FA for six minutes before switching with an Acclaim Pepmap RSLC analytical column (Dinoex-C18, 100 Å, 75 µm × 50 cm). The separation of peptides was performing using a nonlinear 125-minute gradient of solvent A (5% dimethyl sulphoxide (DMSO), 0.1% FA) and solvent B (5% DMSO, 100% ACN, 0.1% FA). The flow gradient was (i) 0-6 min at 3% solvent B, (ii) 6-95 min at 3-23% solvent B, (iii) 95-105 min at 23-40% solvent B, (iv) 105-110 min at 40-80% solvent B, (v) 110-115 min at 80-80% solvent B, (vi) 115-116 min at 80-3% solvent B and equilibrated at 3% solvent B for 10 minutes before the next injection. Briefly, the data was collected using positive polarity with a MS1 scan range of 375-1500 m/z and resolution set to 120,000. Other MS1 instrument parameters include: ACG target of 4e5, maximum injection time of 50 ms and Isolation window of 1.6. MS2 instrument parameters include: scan range of 150-2000 m/z, resolution of 15,000, HCD collision energy of 30%, ACG target of 5e3, maximum injection time of 22 ns and dynamic exclusion of 30 s.

### Sample Preparation of HEK293T, PBMC and Fibroblast Cell Lines

Whole cell pellets of HEK293T cells, PBMCs and fibroblasts were collected and washed twice with PBS before resuspension in 5% SDS, 50 mM tetraethylammonium bromide (TEAB) pH 8.5 buffer and 125 U of benzonase per ml (Sigma-Aldrich). Skeletal muscle samples were solubilized with a probe sonicator with 30% amplitude in 5% SDS, 50 mM TEAB pH 8.5 buffer on ice with a cycle of 10 seconds on 10 seconds off for 1 minute and clarified at 16,000 x g for 5 minutes. Protein concentration was performed with Pierce BCA Protein Assay Kit (Thermo Fisher Scientific) and 25 µg of each sample was aliquoted in triplicates for patients and singlicate for individual controls (N=3-5). Samples were processed using S-trap^TM^ micro spin columns (ProtiFi) according to the manufacturer’s instructions. Proteins were digested with trypsin (Thermo Fisher Scientific) at 1:10 trypsin to protein ratio. Peptides were dried down using a CentriVap Benchtop Vacuum Concentrator (Labconco) and reconstituted in 45 µL 2% ACN, 0.1% trifluoroacetic acid (TFA) and 2 µL injected for liquid chromatography tandem mass spectrometry (LC-MS/MS). Samples were analyzed for 125 minutes (HEK293T) or 95 minutes (Fibroblasts, skeletal muscle, PBMCs) on an Orbitrap Eclipse mass spectrometer (Thermo Fisher Scientific) operating in DIA mode.

### Mass Spectrometry Data Independent Acquisition

Peptides were reconstituted in 45 µL of 2% ACN and 0.1% TFA and 2 µl was injected into an Orbitrap Eclipse mass spectrometer (Thermo Fischer Scientific) for LC-MS/MS analysis equipped with trap and analytical columns described above. All samples were analysed with the mass spectrometer operating in DIA mode. The separation of peptides was performed using a nonlinear gradient of solvent A and solvent B across 125-minutes for HEK293T knockout cell lines, and 95-minutes for all other sample types (fibroblasts, PBMCs and skeletal muscle). Briefly, for the 125-minute method, the flow gradient was (i) 0-6 min at 3% solvent B, (ii) 6-95 min at 3-23% solvent B, (iii) 95-105 min at 23-40% solvent B, (iv) 105-110 min at 40-80% solvent B, (v) 110-115 min at 80-80% solvent B, (vi) 115-116 min at 80-3% solvent B and equilibrated at 3% solvent B for 10 minutes before the next injection. The data was collected using positive polarity with a MS1 scan range of 350-1200 m/z at a resolution of 120,000. Other MS1 instrument parameters include: ACG target of 1e6, maximum injection time of 50 ms, Isolation window of 24 with 1 m/z overlap. For MS2 parameters, the scan range was 200-2000 m/z at 15,000 resolution, HCD collision energy of 30%, AGC target of 1e6 and maximum injection time of 22 ms. For the 95-minute method, the flow gradient was (i) 0-6 min at 3% solvent B, (ii) 6-7 min at 3-4% solvent B, (iii) 7-82 min at 4-25% solvent B, (iv) 82-86 min at 25-40% solvent B, (v) 86-87 min at 40-80% solvent B, (vi) 87-90 min at 80-80% solvent B, (vii) 90-91 min at 80-3% solvent B and equilibrated at 3% solvent B for 5 minutes before the next injection. Instrument parameters were consistent with those used in the 125-minute method described above, with changes made to the following parameters: MS1 scan range of 350-1400 m/z, MS1 maximum injection time of 45 ms, Isolation window of 13.7 and MS2 ACG target of 5e5.

### Proteomic data search

For spectral library, raw files were imported into Spectronaut^®79^ (v.15.2.210819.50606 for HEK293T library, v.14.8.201029.4778 for fibroblasts library, v. 16.0.220606.53000 for PBMCs library) and the three libraries were generated using the ‘Pulsar’ option with default BGS Factory settings, searching against Uniprot human database containing reviewed canonical and isoforms sequences (42,386 entries). The resulting HEK293T library contained 194,270 precursors, the fibroblast library contained 131,627 precursors, and the PBMC library contained 148,522 precursors. For DIA samples, raw files were imported into Spectronaut^®^ and searched against the libraries generated above. Default BGS Factory search parameters were used with changes made to exclude single hit proteins, no selection of ‘Major Group Top N’ and ‘Minor Group Top N’ and data filtering setting set to ‘Q-value’ or ‘Q-value sparse’ with ‘run-wise imputation’ as an imputing strategy. ‘Q-value’ datasets (unimputed) were used for generation of RCA values while ‘Q-value sparse’ with ‘run-wise imputation’ datasets were used to generate heatmaps and correlation plots to reduce missing values for visualization, and in volcano plots for VC and KC cohorts or when the protein of interest was not well detected. UDP1 (*MT-ATP6*) data was also searched using BGS Factory direct DIA settings (library free) with changes as above with inclusion of the p.Met228 variant peptide fasta file in the protein database search. SC1 and SC2 (*TMEM70*) skeletal muscle were searched with default BGS Factory direct DIA settings with changes as above.

### Protein filtering and clean up

Proteins were filtered in each cohort to ensure that the data was of high quality and with a minimal amount of missing data for analysis. Proteins within the validation cohort (VC) were filtered out if identified by a single peptide in all controls or proband samples, maintaining proteins that were identified by >2 peptides in both groups. Proteins were then filtered to least three or more valid values for the controls and at least two or more valid values for the proband samples. For the knockout cohort (KC), proteins were filtered out if identified by a single peptide in all controls and proband samples. Proteins were then filtered to at least two valid values for the controls and at least two valid values for the knockout sample. For the undiagnosed proband cohort (UDP), proteins were filtered out if identified by a single peptide in all controls or proband samples. Proteins were then filtered for 70% valid values for the controls due to the different number of controls per batch and 70% valid values for the proband samples. For the supporting cohort (SC), samples SC1, SC2, SC3 and SC5 proteins were filtered out if identified by a single peptide in all controls or proband samples. Proteins were then filtered for 70% valid values for the controls and 70% valid values for the proband samples. Sample SC4 had no filtering completed as this was a previously published dataset^21^ and the data was unlogged and complex subunit gene names manually updated to match the current MitoCarta3.0 naming convention for Complex V subunits. For the 36 PBMC control samples (SC6-41), proteins were filtered out if they were identified by a single peptide across all 36 samples and further filtered for at least 70% valid values.

### Relative Complex Abundance Plots

Relative complex abundance (RCA) plots were generated for all VC, KC, SC and UDP cohorts and plots can be viewed within the RDMS Explorer platform. All samples underwent mitochondrial normalization where the mean of the mitochondrial proteins for each specific sample was determined using the average of all control mitochondrial proteins or proband mitochondrial proteins and then used as a normalization factor for each group and applied to each sample. To develop the RCA plots, the average of the proband and controls was determined and then transformed to log10. The difference between the log10 of the proband and controls was determined and then unlogged. Statistical difference between the control and proband complex subunits was determined using a paired t-test and summary statistics including mean and the error bars indicating standard deviation were determined for each complex and the graph plotted within R (v.4.3.2). Complex subunits were determined using curated annotations from MitoCarta3.0^28^. For Supplementary Fig. A, relative mitochondrial abundance from MitoCarta3.0 ‘+’ entries were generated as above without the mitochondrial normalization step. For Supplementary Fig. B, control mitochondrial abundance plots were generated using log2 abundances from MitoCarta3.0 ‘+’ entries for each batch and significance performed with an ANOVA test.

### Protein range graphs

Protein range graphs were developed using an in-house R script. The median of the protein of interest in control samples is calculated as well as the median of the proband replicates. The log2 fold-change is calculated between the proband and control and converted to percentage for the proband as the abundance relative to the control median for plotting. The range of the controls is also displayed as the abundance relative to the control median. The log2 abundance per standard deviation (SD) is rounded to one decimal place with the top axis SD present to determine how many standard deviations the protein is from the control median.

### Volcano and Correlation plots

The peptide and valid value cleaned matrices for all samples were annotated with MitoCarta3.0^28^ entries in Perseus^80^. All analyses were performed in Perseus v 1.6.14.0. Volcano plots were visualized using scatter plot function from a two-sample t-test using p-value threshold of 0.05 in Perseus. For the correlation plots (Fig 3B, Supplementary Fig.2B), proteomic data searched with q-value sparse with run-wise imputation settings were correlated via gene name using gene name entries and the logFCs in R and plotted using scatter plot function in Perseus. For Fig. 4C, previously published data from MRPL50 Fibroblasts^21^ were correlated with MRPL50 PBMCs using gene name entries and the logFCs in R and plotted using scatter plot function in Perseus.

### RDMS Explorer Website

All the code used to develop the RDMS Explorer website (https://rdmassspec.shinyapps.io/RDMSExplorer/) and in-house analysis performed in R can be found at (https://doi.org/10.5281/zenodo.12883602) and (https://github.com/njcaruana/rdms_oxphos).

### Heatmaps and Venn diagrams

For heatmaps output matrices searched with q-value sparse run-wise imputation settings Spectronaut runs were log_2_ transformed and a two-sided t-test using p-value for truncation was performed in Perseus for each cell line against their respective batch controls. The Morpheus software was used for heatmap visualization (https://software.broadinstitute.org/morpheus). For Supplementary Fig. 1A, proteins were manually annotated based on MitoCarta3.0 as ‘assembly factor’ or ‘complex I-V’ and subset for visualization in Morpheus using the following parameters metric ‘one minus Pearson correlation’, linkage method ‘average’, cluster ‘rows’ and group columns by ‘complex/assembly factor’. P-value results from t-test statistical analysis were used to visualize the significance of each data point by overlaying the p-value matrix onto the t-test matrix in Morpheus and using the option size by ‘p-value matrix’ size minimum ‘0’ and maximum ‘1.301’ (p-value<0.05). For the correlation heatmap plot (Supplementary Fig. 2A), hierarchical clustering was used with the following parameters in Morpheus: metric ‘one minus Pearson correlation’, linkage method ‘average’ and cluster ‘columns’. Topographical heatmaps (Supplementary Fig. 1B) were generated from log_2_-transformed fold-change from t-test in Pymol using the PDB structures (CI: 5LDW, CII: 1ZOY, CIII: 1BGY, CIV: 5Z62, CV: 7AJD) as previously described^26^. Venn diagrams were generated with DeepVenn^81^ from gene names. Green (diagnostic-grade) and amber (borderline diagnostic-grade) lists for mitochondrial disease (v.0.787) and Mendeliome gene lists (v.0.12869) were retrieved from PanelApp Australia^27^.

### Principal Component Analysis and Donut plots

PCA plots were generated in Perseus v 1.6.14.0 from t-test fold-change data for VC cohort (Fig. 1B) or protein abundances for PBMCs controls (Fig. 4B). Donut plots were generated in GraphPad Prism (v.10.2.0).

### RNA sequencing and transcriptomic analysis

RNA sequencing and analysis was performed as described^20^. In brief, RNA was extracted from muscle using miRNeasy Mini Kit (Qiagen) and treated with RNase-Free DNase (Qiagen) and RNA quality and quantity measured using TapeStation RNA ScreenTape analysis (Agilent) and Qubit RNA HS (Thermo Fisher Scientific). UDP4 underwent RNAseq within a group of 10 undiagnosed patients with available muscle samples, all with an RNA integrity number of 7.2 - 8.9. The Kapa Biosystems mRNA Hyper Prep Kit was used to generate sequencing libraries from 500ng RNA, and paired-end sequencing on an Illumina instrument was performed at the Yale Center of Genome Analysis to achieve 50-100 million read coverage.

For outlier detection, 100 control muscle samples were retrieved from GTEx using the Sequence Read Archive (SRA) toolkit and converted into FASTQ using Fastq-dump. Patient and control FASTQ files were aligned to hg38 human reference genome with GENCODE v26 annotations using STAR (v2.5.3a), with twopassMode = ‘Basic’ to enable detection of novel splice junctions, and duplicates marked using Picard (v2.9). RNA-SeQC (v2.3.4) was used to generate quality metrics from STAR-aligned bam files. Outliers were removed per the following criteria: (a) <45 million QC-passed reads, (b) <18,000 genes detected and (c) manual identification via Principal Component Analysis. This resulted in the removal of 11 samples, leaving 89 control samples from muscle. Outlier expression was assessed using the DROP (Detection of RNA Outliers Pipeline) pipeline v0.9.0 (https://github.com/gagneurlab/drop), with default settings^82^. Analysis was restricted to 6106 genes, which included Mendeliome and ‘MitoExome’ genes, as described previously^20^.

### cDNA studies

The total RNA extracted from skeletal muscle for RNA sequencing was also synthesized into cDNA using the SuperScript III First-Strand Synthesis System (Thermo Fisher Scientific) as per both manufacturer’s protocols and as described previously^83^. To examine mRNA splicing in *CCDC47*, PCR primers (5’ GCGTGACTGAGCTACGGTT 3’; 5’ CTCTGGGATGGCTTTACATGG 3’) were designed to amplify the entire ORF from cDNA. PCR products were analyzed on 1% agarose (Bioline) gels using a 1-kb Plus DNA Ladder (Thermo Fisher Scientific) prior to cloning into a pCR2.1-TOPO vector using the TOPO TA cloning kit (Thermo Fisher Scientific) and transformation into TOP10 competent cells (Thermo Fisher Scientific). Individual colonies were examined and then underwent sanger sequencing.

## Data Availability

The mass spectrometry proteomics data have been deposited to the ProteomeXchange Consortium via the PRIDE84 partner repository with the dataset identifier PXD055473. De-identified genomic data from this study are available for ethically approved research. For queries about the genomic data sets please contact: . All the code used to develop the RDMS Explorer website (https://rdmassspec.shinyapps.io/RDMSExplorer/) and in-house analysis performed in R can be found at (https://doi.org/10.5281/zenodo.12883602) and (https://github.com/njcaruana/rdms_oxphos).

## MitoMDT Diagnostic Network for Genomics and Omics

David R Thorburn, Aleksandra Filipovska, Michael T Ryan, David A Stroud, Diana Stojanovski, David Coman, Sean Murray, Ryan L Davis, John Christodoulou, Roula Ghaoui, Suzanne CEH Sallevelt, Cas Simons, Stefan J Siira, Shanti Balasubramaniam, Alison G Compton, Daniel G MacArthur, Nicole J Lake, Amanda Samarasinghe, Yoni Elbaum, Catherine Atthow, Pauline McGrath, Ellenore M Martin, Madeleine Harris, Tegan Stait, Leah E Frajman, Simone Tregoning

## Author contribution

Conceptualization: DHH, DRT, DAS

Methodology: DHH, NJC, LNS, NJL, LuEF, SSCA, TS, ST, LeEF, DRLR, MB, BoR, AGC, VK, CA

Investigation: DHH, MB, BrR, MJW, AV, CB, HP, JL, ZS, RM, RWT, AGC, WB, NBT, MLF

Visualization: DHH, NJC, LNS, NJL, SSCA

Website design and coding: NJC

Funding acquisition: DHH, JC, AGC, DRT, DAS

Project administration: AS, RB, DRT, DAS

Supervision: PM, ML, RM, RWT, MTR, ZS, JC, AGC, DRT, DAS

Writing – original draft: DHH, DRT, DAS

Writing – review & editing: all authors reviewed and edited the manuscript

## Acknowledgments

We acknowledge and thank all the families who contributed to this study. We thank all members of the Stroud and Stojanovski lab for input into experimental design and interpretation of data. We thank the Bio21 Mass Spectrometry and Proteomics Facility (MMSPF) for the provision of instrumentation, training, and technical support. The Chair in Genomic Medicine awarded to JC is generously supported by The Royal Children’s Hospital Foundation. The authors thank staff of the Pathology Collection Department at The Royal Children’s Hospital for obtaining the consent of participants and the collection of samples. The authors thank staff of the Anaesthetic and Surgical Departments at the Royal Children’s Hospital.

## Data Availability Statement

The mass spectrometry proteomics data have been deposited to the ProteomeXchange Consortium via the PRIDE^84^ partner repository with the dataset identifier PXD055473. De-identified genomic data from this study are available for ethically approved research. For queries about the genomic data sets please contact.

