## Supplementary Material 1 for "Untargeted proteomics enables ultra-rapid variant prioritization in mitochondrial and other rare diseases"

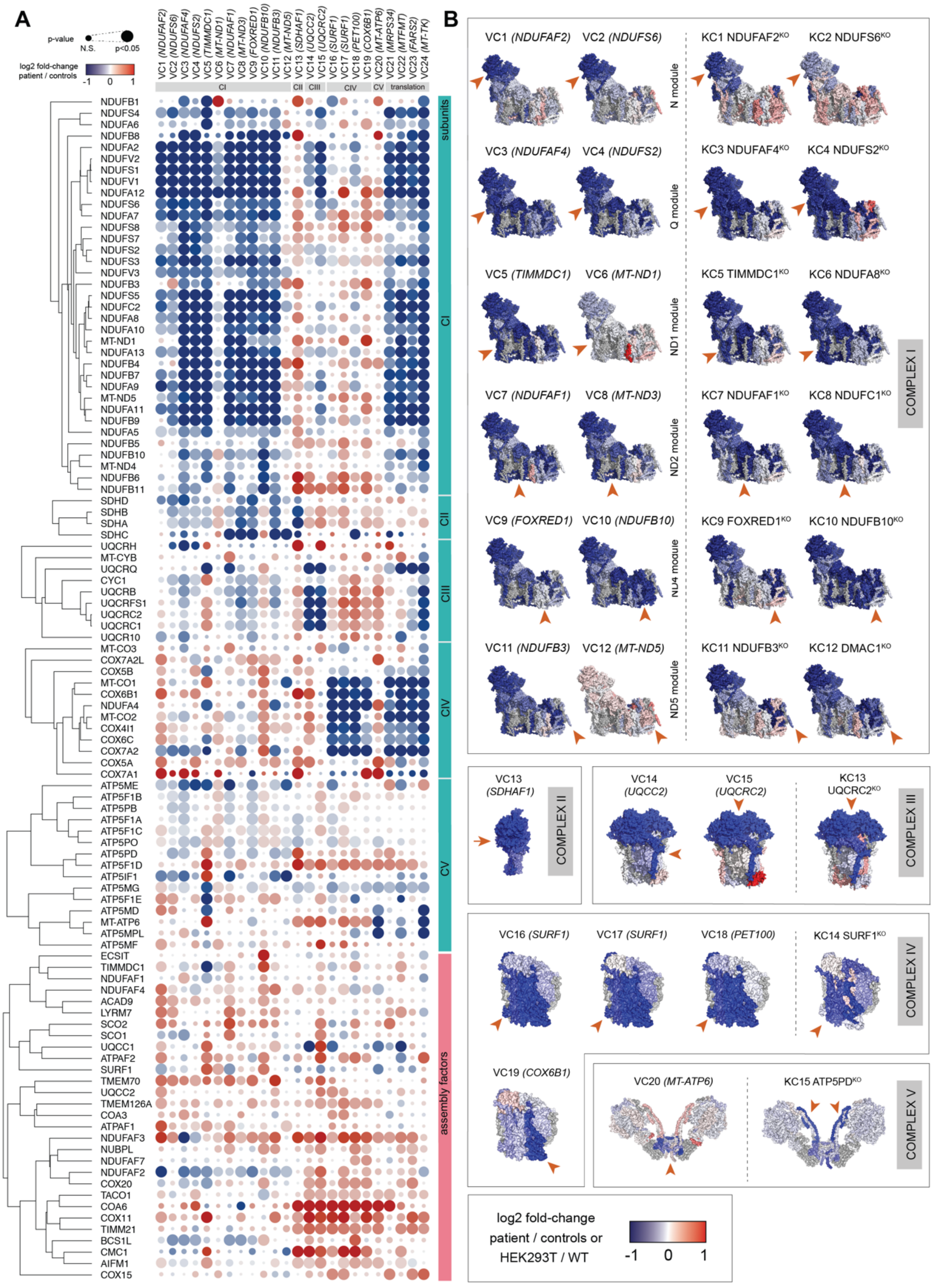

**Supplementary Figure 1**. **A –** Heatmap and hierarchical clustering of OXPHOS subunits by complex and assembly factors in the validation cohort (VC) using fold-change values calculated using t-test analysis from baseline imputed proteomic dataset. **B –** Topographical heatmap using fold-change values calculated from t-test analysis using baseline imputed proteomic dataset for isolated defects in the validation cohort (VC). PDB ID: 5LDW (CI), 1ZOY (CII), 1BGY (CIII), 5Z62 (CIV), 7AJD (CV).

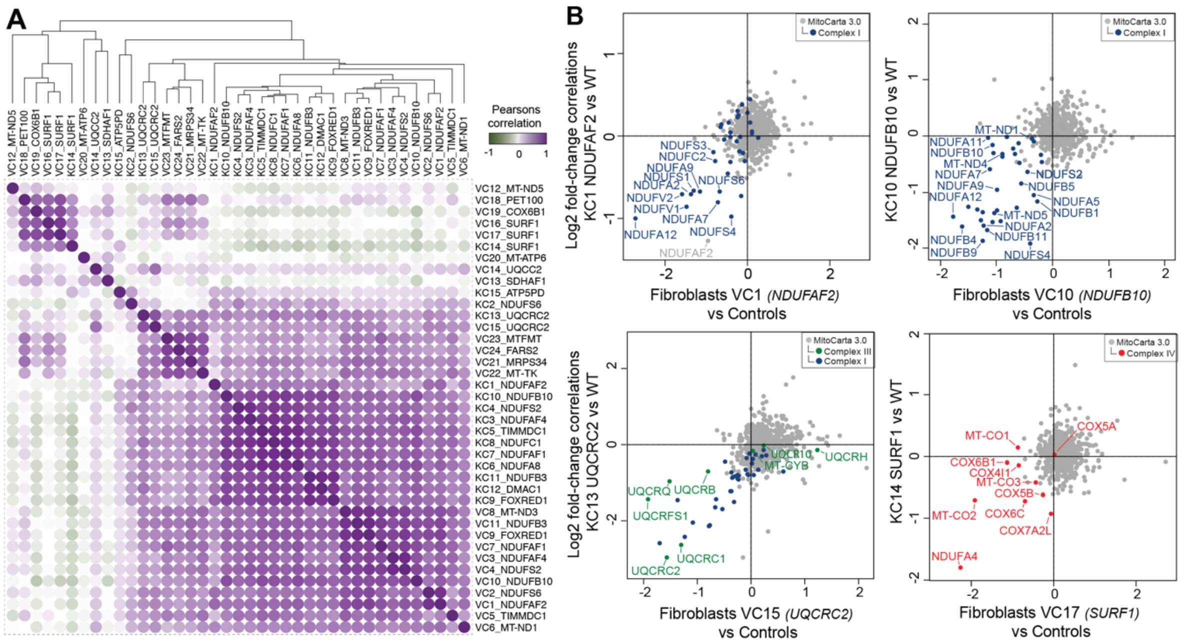


**Supplementary Figure 2**. **A –** Pearson correlation between validation cohort (VC) and knockout cohort (KC) using fold-change values for OXPHOS subunits annotated with MitoCarta3.0 calculated from t-test analysis using baseline imputed proteomic dataset. **B –** Pairwise correlation of fibroblast line from validation cohort (VC) and equivalent HEK293T knockout cohort (KC) from baseline imputed proteomic dataset for mitochondrial proteins highlighting the expected affected complex.

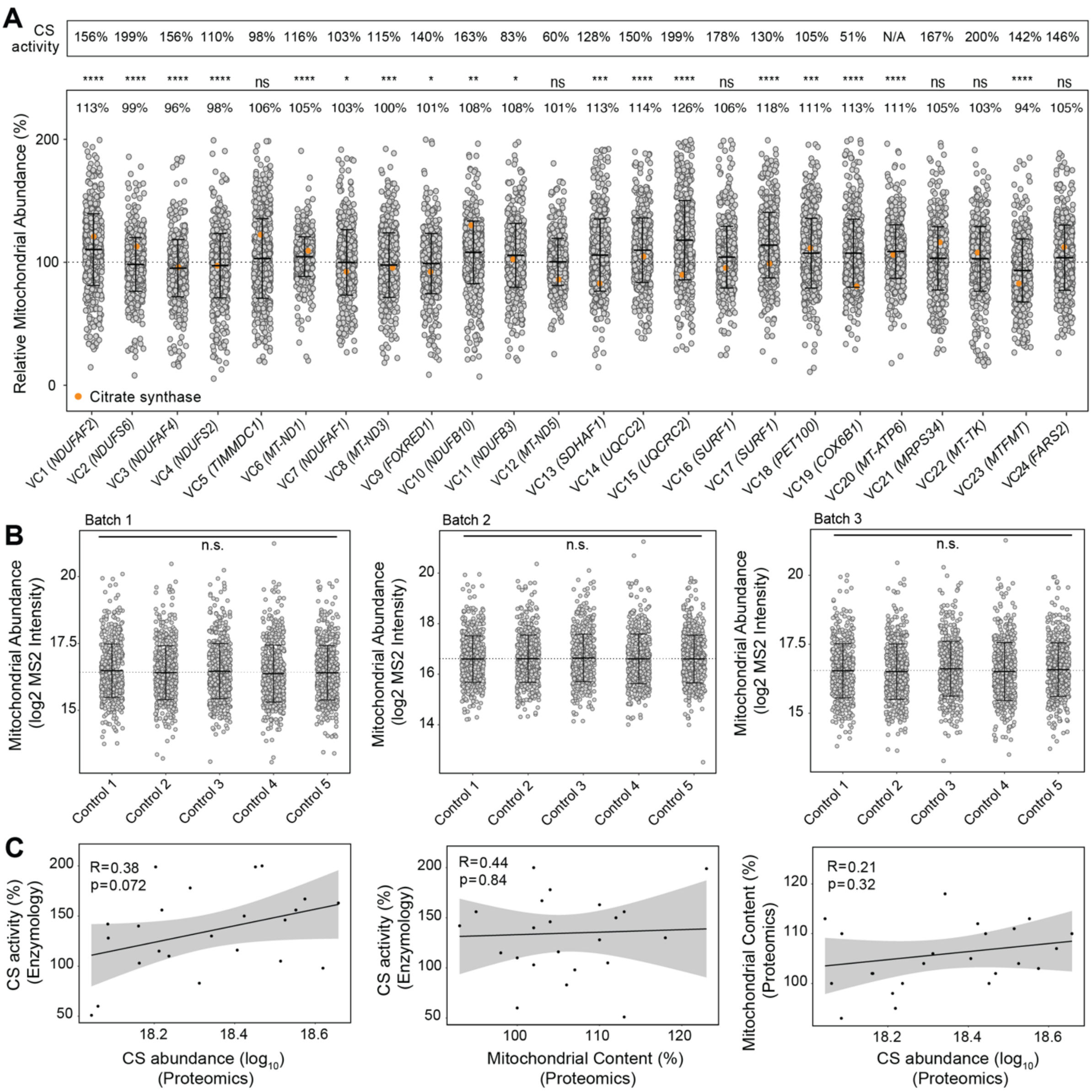


**Supplementary Figure 3**. **A –** Citrate synthase (CS) activity of fibroblast lines relative to controls and relative abundance of mitochondrial proteins (MitoCarta3.0) for the validation cohort (VC) compared to controls depicting variable abundance of mitochondrial levels across cell lines. Middle bar represents mean mitochondrial abundance. Upper and lower bars represent 95% confidence interval. Significance was calculated from a paired t-test between the mitochondrial means. Orange dot = citrate synthase protein abundance. **** = p<0.0001, *** = p<0.001, ** = p<0.01, * = p<0.05, ns = not significant, *p*>0.05. **B –** Relative abundance of mitochondrial proteins (MitoCarta3.0) in the five controls used in each batch of the validation cohort (VC) showing no significant changes using an ANOVA test. **C –** Pearson correlation between CS activity from enzymology and CS abundance from proteomics (left), CS activity from enzymology and mitochondrial content from proteomics (middle); and mitochondrial content from proteomics and CS abundance from proteomics (right).

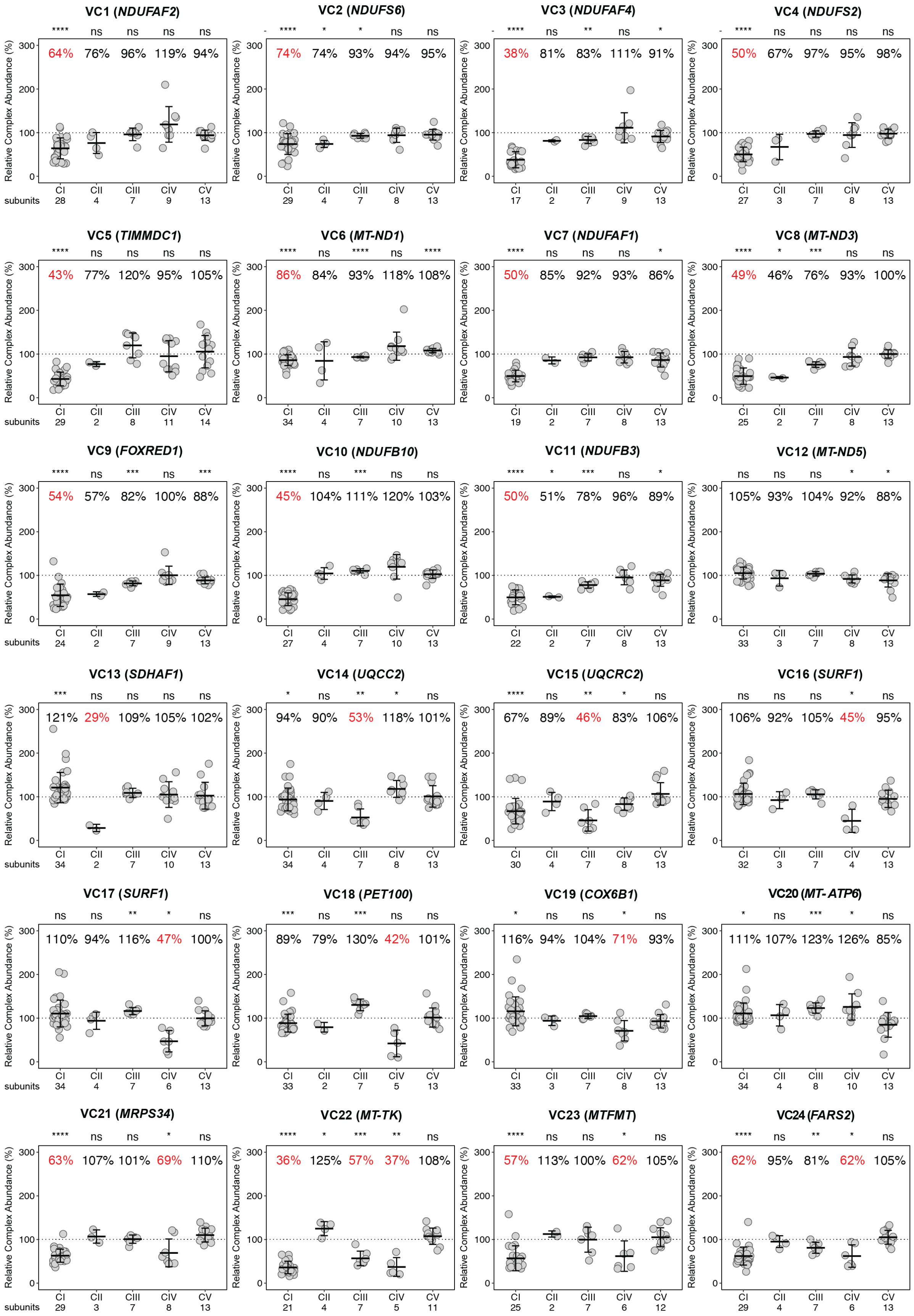

**Supplementary Figure 4.** Relative Complex Abundance (RCA) of OXPHOS complexes in the validation cohort (VC) from non-imputed proteomic dataset. Middle bar represents mean complex abundance. Upper and lower bars represent 95% confidence interval. Significance was calculated from a paired t-test between the individual protein means. **** = p<0.0001, *** = p<0.001, ** = p<0.01, * = p<0.05, ns = not significant, $p$>0.05.

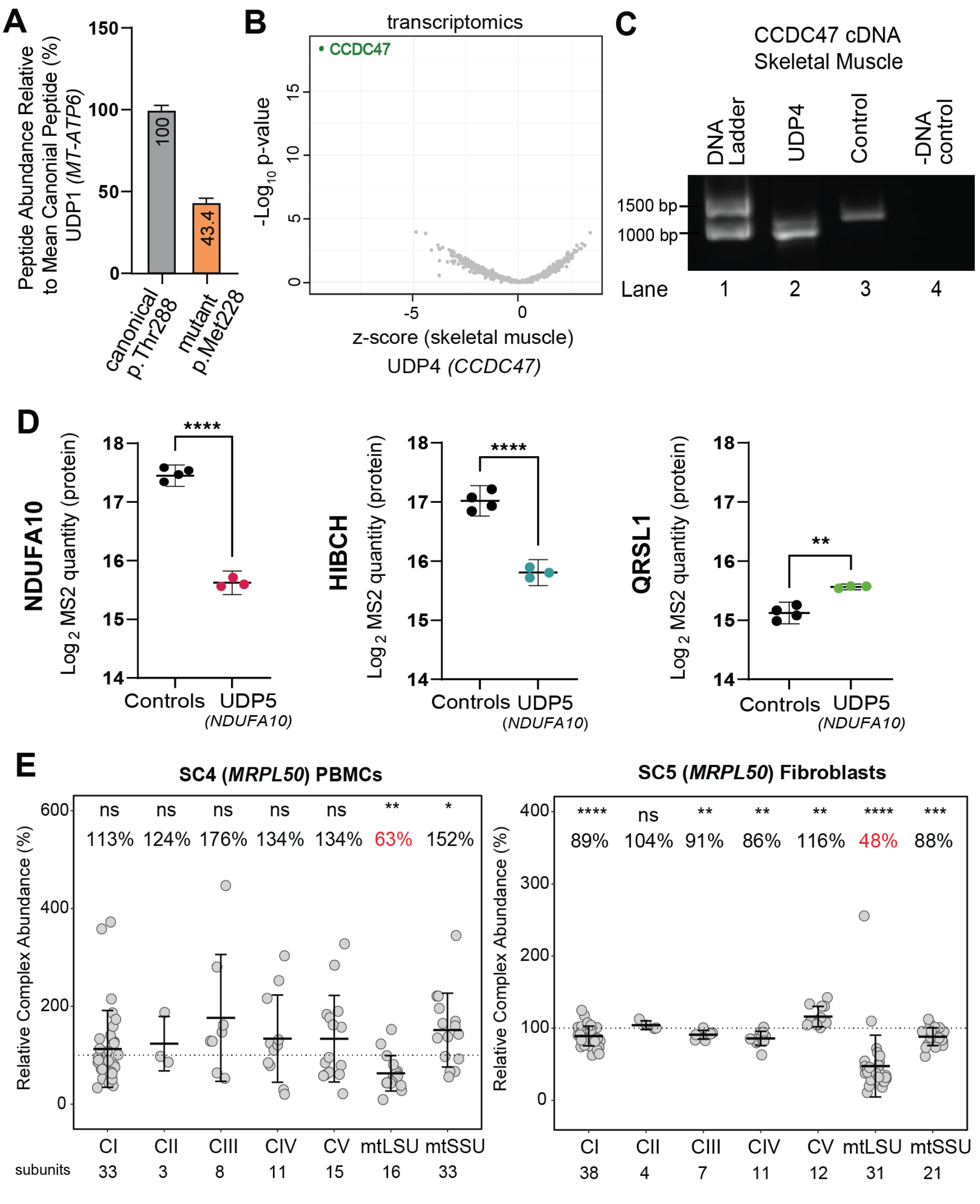


**Supplementary Figure 5**. **A –** Abundance of the ATAD3A mutant peptide (p.Met288) relative to the canonical peptide (p.Thr288). **B –** Transcriptomic analysis of skeletal muscle for UDP4 (*CCDC47*) relative to controls showing strong reduction of *CCDC47* transcripts. **C –** cDNA analysis of UDP4 (*CCDC47*) using RNA extracted from skeletal muscle. Gel electrophoresis of full-length PCR products designed to amplify the entire open reading frame (ORF) show the presence of both missense transcripts and a second shorter one, missing 121 bp from the 3' end of exon 4 from a donor-splice site five nucleotides upstream of the missense variant. **D –** Relative abundance of NDUFA10, HIBCH and

QRLS1 proteins from whole cell fibroblasts in UDP5 (*NDUFA10*) compared to controls. Significance was calculated from an unpaired t-test between the means. **** = p<0.0001, ** = p<0.01. **E –** Relative Complex Abundance (RCA) of OXPHOS complexes and mitoribosome large (mtLSU) and small (mtSSU) subunits in *MRPL50* peripheral blood mononuclear cells (PBMCs)(left) and fibroblasts (right). Middle bar represents mean complex abundance. Upper and lower bars represent 95% confidence interval. Significance was calculated from a paired t-test between the individual protein means. **** = p<0.0001, *** = p<0.001, ** = p<0.01, * = p<0.05, ns = not significant, *p*>0.05.
